# Mortality, morbidity, and post-operative complications of typhoid intestinal perforations: global systematic review and meta-analysis

**DOI:** 10.1101/2024.06.28.24309663

**Authors:** Nienke N. Hagedoorn, Megan Birkhold, Shruti Murthy, Suzanne Faigan, Meera D. Rathan, Christian S. Marchello, John A. Crump

## Abstract

**Background:** TIP is a serious and life-threatening complication of typhoid fever that requires emergency surgery and an important driver of typhoid burden. We aimed to review global studies reporting on mortality, morbidity, and post-operative complications in patients with typhoid intestinal perforation (TIP).

**Methods:** We searched multiple databases for articles reporting case-fatality ratio (CFR) or complications in patients with TIP undergoing surgery published from 1980 through 11 January 2025. We described the prevalence of each reported complication. Of patients with TIP, we pooled CFR using random-effects meta-analysis and stratified by United Nations region, sex, and number of perforations per patient.

**Results:** We included 48 articles reporting on 4,309 patients with TIP. The most prevalent post-operative complications were wound or surgical site infection in 1,553 (50.1%) of 3,100 patients, wound dehiscence in 308 (16.1%) of 1,909, and respiratory infection in 136 (15.6%) of 872. Overall, the pooled CFR (95%CI) of patients with TIP was 16.3% (13.4-19.7%), and was 20.3% (17.0-24.2%) in 32 observations from the African region, 8.5% (5.7-12.6%) in 15 observations from the Asian region. Overall, no significant correlation was observed between median year of data collection and CFR (estimate [95%CI]: 0.00 [-0.02, 0.02]), nor when analyses were stratified by region, including Africa (estimate [95%CI]: −0.01 [-0.03, 0.02]), and Asia (estimate [95%CI]: −0.04 [-0.07,0.00]).

**Conclusions:** Disability and death associated with TIP remains substantial. Over time, mortality from TIP did not decline. Efforts to improve access to and quality of surgical services for those with TIP are warranted.

## Introduction

Typhoid fever is caused by systemic infection of *Salmonella enterica* subspecies *enterica* serovar Typhi (*Salmonella* Typhi). The clinical diagnosis of typhoid fever is challenging as presenting symptoms such as fever, malaise, and headache are similar to those of other febrile illnesses.[1] Delays in diagnosis and in receipt of appropriate antimicrobial treatment are associated with increased risk for serious complications, including gastrointestinal bleeding or typhoid intestinal perforation (TIP).[2, 3] While gastrointestinal bleeding can usually be managed non-surgically, TIP is a serious and life-threatening complication that requires emergency surgery.

In hospitalized patients with typhoid fever, the prevalence of TIP was estimated to range from 1 to 3%.[4–7] Among TIP patients, 52-92% occur in those aged <30 years.[8, 9] Additionally, some TIP studies found a male preponderance,[8, 10, 11] while others did not.[9, 12] The case fatality ratio (CFR) of TIP is high, estimated approximately 40% in studies published from 1960 through 1990.[6, 7] In more recent studies, mortality was approximately 15%, attributed in part to improvements in pre- and post-operative management and surgical techniques.[4, 11, 13] Clinical predictors of mortality in TIP patients include the presence of shock and of multiple perforations.[8]

Patients with TIP also experience high morbidity. Common complications following surgery include surgical site infections, pneumonia, re-perforation, and enterocutaneous fistula formation.[13, 14] Birkhold et al performed a scoping review of morbidity following TIP surgery including data from countries in sub-Saharan Africa, and found that the prevalence of complications among studies ranged from 16% to 100%. Surgical site infections occurred in more than 60% of patients having surgery for TIP among surgical centers in 11 countries.[15]

Despite being an important consideration for burden of disease estimation, previous systematic reviews of post-operative complications of TIP have focused on specific regions, countries, or age groups.[11, 13, 16–18] We aimed to perform a systematic review to synthesize global data on mortality, morbidity, and post-operative complications in patients with TIP.

## Methods

### Study design and search strategy

We performed a subanalysis of the systematic review and meta-analysis on complications and mortality of patients with typhoid fever by Murthy *et al*.[19] In this subanalysis (PROSPERO CRD42023438367),[20] we included ‘surgical’ observational study designs or control arms of clinical trials that reported mortality, morbidity, or post-operative complications in patients undergoing surgery for TIP.

PubMed, Web of Science, and pre-prints servers and repositories were searched to identify articles on typhoid fever complications and mortality published from 1 January 1980 through 11 January 2025 in any language. We excluded articles that did not describe the criteria of classifying a case of TIP, based TIP case definitions on serology or clinical symptoms, or used culture of a non-sterile site to isolate *Salmonella* Typhi. We excluded case reports, policy reports, conference abstracts, and government passive surveillance reports. In addition, we excluded articles that provided only summary data or provided no clear denominator to calculate proportions. Additional eligible articles were identified by reviewing reference lists of eligible articles. The study was reported according to the Preferred Reporting Items for Systematic Reviews and Meta Analyses (PRISMA) guidelines (Supplementary Materials 1).[21]

A case of ‘confirmed TIP’ was defined as a perforation in a person confirmed by histopathology stains using *Salmonella* Typhi-specific immunohistochemistry, or a positive blood culture for *Salmonella* Typhi to attribute the cause of perforation to *Salmonella* Typhi. A case of ‘probable TIP’ was defined as a perforation in a person identified by gross intraoperative findings that included any of the following terms: ‘terminal ileum,’ ‘antimesenteric perforation,’ or ‘based on gross appearance at laparotomy.’[13, 22]

Records were deduplicated in EndNote (Clarivate, London, United Kingdom). Subsequently, references were uploaded to Rayyan (Qatar Computing Research Institute, Dohar, Qatar) for title-and-abstract and full-text screening. Study selection and data abstraction were performed independently by two reviewers (any of CSM, MB, NNH, SF, or SM). Discrepancies were resolved by discussion or by a third reviewer (JAC).

### Data abstraction

We abstracted the following data: publication year, data collection period, study design, country, hospital level (district or rural hospital, regional hospital, university hospital),[23] availability of critical care services, duration of follow-up of participants to ascertain outcomes and complications, quality of care delivered to patients with TIP (e.g., use of antimicrobials, fluid resuscitation, blood transfusion, urine output monitoring, nasogastric tubes, parenteral nutrition), American Society of Anesthesiologists (ASA) Physical Status Classification System by individual patient, age range and sex, number of patients with severe peritoneal contamination defined as ‘severe,’ ‘gross,’ ‘fecal,’ or ‘>1000 mL pus’; number of TIP deaths, stratified by sex and number of perforations (single/ multiple) per patient; surgical technique (primary repair, wedge resection, resection and anastomosis, resection and ostomy creation, hemicolectomy or colectomy, or other).

We abstracted all complications at presentation to the hospital and post-operative complications reported by authors. For complications at presentation, we used a pre-defined list of 21 complications of Parry *et al*.[3] For post-operative complications, we abstracted a pre-defined list based on commonly reported complications identified in a previous review.[24] We abstracted duration-related variables in units reported by the authors of included articles and subsequently converted the units to days: days from illness onset to presentation, days from illness onset to TIP diagnosis, days from illness onset to surgery, days from TIP diagnosis to surgery, days from TIP diagnosis to death, and total length of hospital stay.

We categorized the age of participants into children <16 years, adults ≥16 years, or mixed children and adults. Countries were classified by United Nations (UN) region and subregion.[25] As a proxy for level of development, countries were classified by income level following the 2024 World Bank scheme as low income countries (LICs), lower-middle income countries (LMICs), upper-middle income country (UMICs), and high income countries (HICs).[26]

Risk of bias was assessed following the Joanna Briggs Institute guidance for critical appraisal on prevalence studies.[27, 28] Additional specific questions to assess study design, complications, and follow-up were added (Supplementary Materials 3).

### Data analysis

The proportion of each complication was calculated by dividing the number of patients with each complication by the number of patients in whom the complication was assessed, or by the number of patients with TIP, as applicable. We calculated the number of patients with complications at presentation, and for post-operative complications. For post-operative complications that were reported in ≥100 patients for each UN region, we compared prevalence by UN region using the Chi^2^ test. We additionally reported the morbidity ratio. The morbidity ratio was defined as the number of patients with any complication divided by all patients with TIP. As proxy for more advanced disease, we assessed the proportion of patients with multiple perforations and for severe peritoneal contamination. We reported the median (IQR) of mean duration from symptom onset to presentation, symptom onset to perforation, and length of hospital stay.

We assessed the CFR by dividing the number of deaths by number of patients with TIP. Among articles, we described the CFR stratified by UN region and World Bank income group [25, 26] sex; age group; and single versus multiple perforations. We assessed the correlation of the median year of data collection with the CFR using Pearson’s rho and stratified per UN region.

We pooled the CFR by performing an inverse variance random-effects meta-analysis with logit transformation with Hartung-Knapp adjustment and Sidik-Jonkman estimator,[29, 30] in R version 4.3.3 (meta package) (R Foundation for Statistical Computing, Vienna, Austria) when two or more estimates for CFR were available. We performed subgroup analyses when ≥10 estimates by subgroup were available by UN region, by age group, by sex, and by number of perforations. A mixed-effects meta-regression was performed to investigate the heterogeneity on TIP CFR of moderators such as country, age group, study quality, income group, ICU and parenteral nutrition availability, and median year of data collection. We assessed and plotted using bubble plots, from the meta-regression, the variation per estimate of typhoid fever CFR, overall and by UN regions, with median year of data collection, by regression coefficient (β) with 95% CI. We assessed heterogeneity among articles was assessed using forest plots, Chi^2^ test, *I^2^* statistic, and Tau^2^ (τ^2^). We considered an I^2^ value of 0–49% as low, 50–74% as moderate, and ≥75% as substantial heterogeneity.[31] We considered a p-value <0.05 as indicating significant heterogeneity. Subgroup differences were assessed using the Chi^2^ test.

To explore differences in mortality among UN regions, we investigated potential risk factors on article-level for mortality and evaluated hospital-level, availability of critical care services, and availability of parenteral nutrition as proxy for available health facility resources. We selected parenteral nutrition among available quality of care items as use of parenteral nutrition requires more specialized equipment and experienced personnel than the other items. Other included article-level risk factors were mean symptom duration until presentation, the proportion of patients with duration of perforation to surgery of ≥24 hours, and the proportion of patients with severe peritoneal contamination. For continuous variables, we assessed the correlation with CFR using Pearson’s rho and for categorical variables the CFR was compared with the Mann-Whitney U test. All data analyses were performed in R version 4.3..

## Results

We included 29 articles from the primary review, and 22 additional articles among 8,850 identified in the updated search. Subsequently, three articles of the primary review were excluded because of overlapping time periods leading to 48 articles for this subanalysis on TIP (see flowchart in Supplementary Materials Figure 1; details of included articles in Supplementary Materials 4).

**Figure 1.**
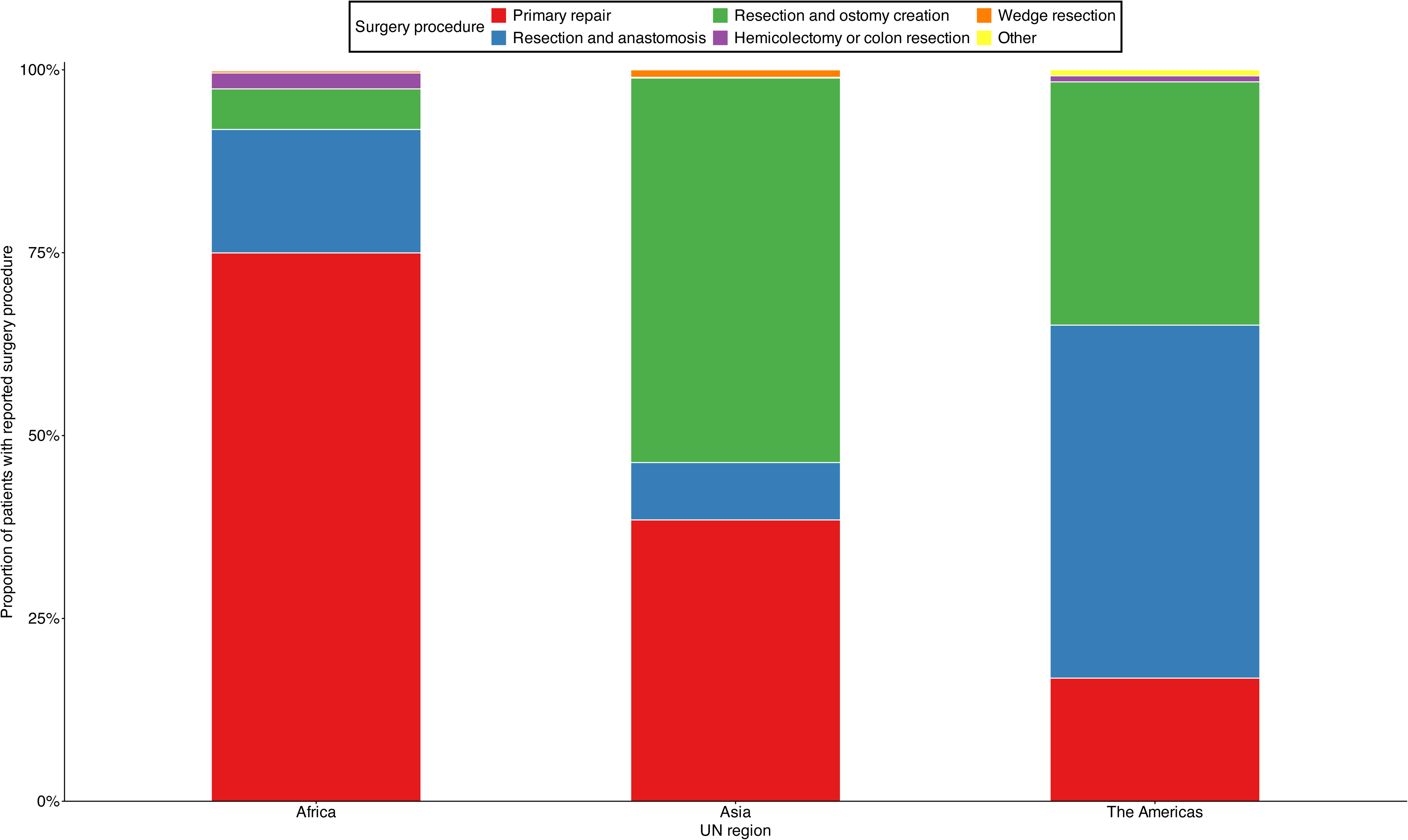
Frequency of surgical procedures by UN region among patients with typhoid intestinal perforations, global systematic review and meta-analysis, 1966-2023

Of 48 articles that collected data 1966-2023 from 62 sites in 23 countries yielded 51 observations, 33 (68.8%) articles reported data from 15 African countries, 12 (25.0%) articles from six Asian countries, and three (6.3%) from two American countries. (Supplementary Materials Figure 2). The studied population was children for 11 (21.5%) observations, adults in five (9.8%) observations, and mixed children and adults in 35 (68.6%) observations.

**Figure 2.**
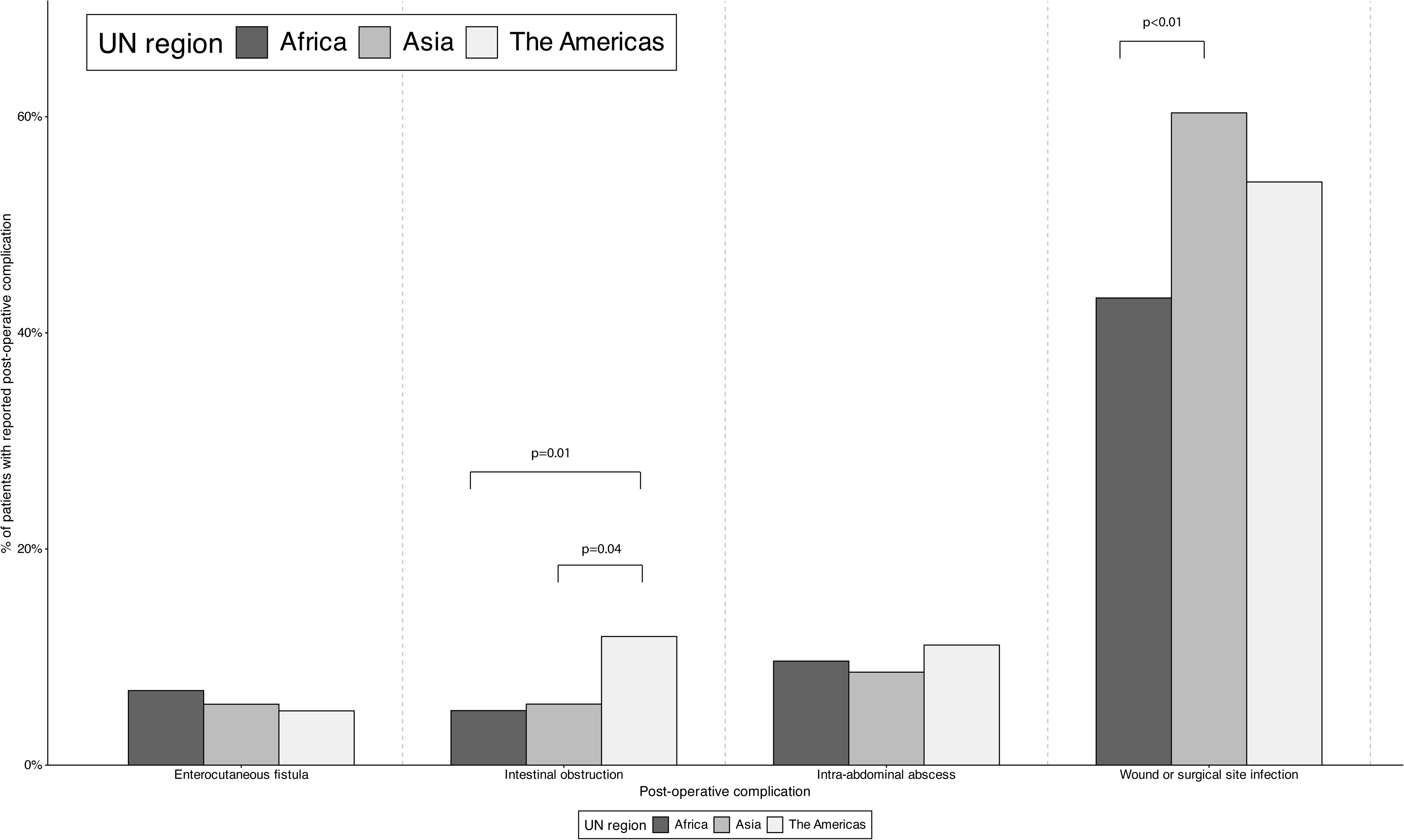
Prevalence of post-operative complications among patients with typhoid intestinal perforation by UN region, global systematic review and meta-analysis, 1966-2023 Legend: Brackets show significant comparisons between UN regions. P-values were adjusted for multiple testing according the Bonferroni method.

Among 35 observations from which the hospital level could be abstracted, 24 (68.6%) were at a university hospital (Supplementary Materials 5). The reported care available to patients was antimicrobial treatment in 36 (75%) articles, fluid resuscitation in 30 (62.5%), blood transfusion in 11 (22.9%), urine output monitoring in 16 (33.3%), nasogastric tubes in 13 (27.1%), and parenteral nutrition in three (6.3%). The ASA physical status of patients was reported in six (12.5%) articles. The overall risk of bias was high for 44 (91.6%) articles, intermediate for two (4.2%) articles, and low for two (4.2%) (Supplementary Materials Figure 3).

**Figure 3.**
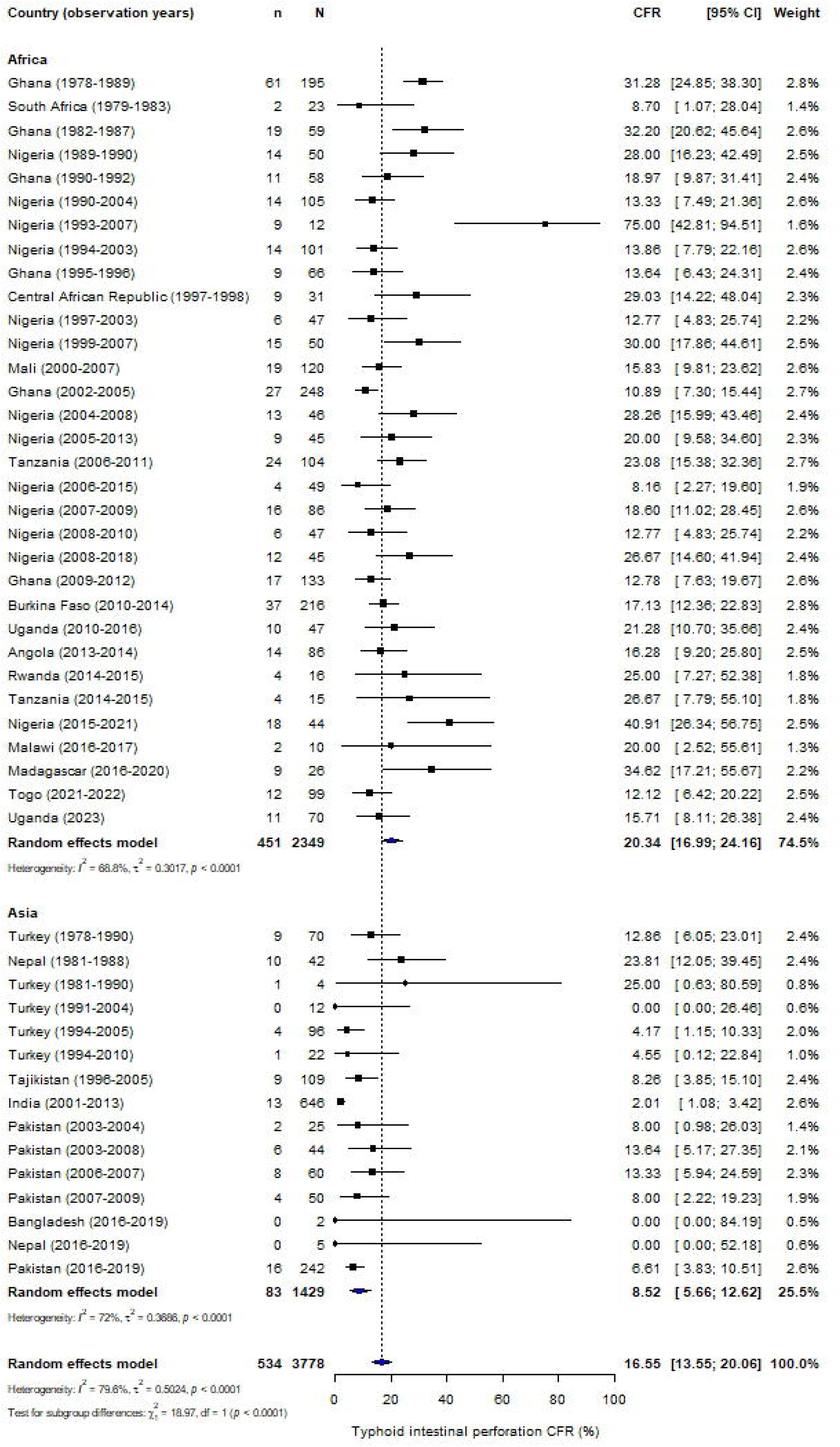
Meta-analysis of case fatality ratio (CFR) in patients with typhoid intestinal perforations per UN region Legend: TIP, typhoid intestinal perforations

Among 51 observations, the median (IQR) number of patients with probable or confirmed TIP was 50 (29-100) per observation. Of 4,309 patients with probable or confirmed TIP, 950 (22.0%) were confirmed TIP (Supplementary Materials 5). Of 4,309 patients, 2,397 (55.6%) were from Africa. Among 42 observations that reported sex, the median (IQR) proportion of males was 71.2% (63.4-76.5%) per observation. Among 34 articles reporting number of perforations among 2,620 patients, the median (IQR) proportion with single perforations was 74.9% (70.2-83.3%) per observation (Supplementary Materials 5). Complications at presentation to the hospital for TIP patients are presented in Supplementary Materials 6.

Surgical procedures were described in 39 (81.3%) articles reporting on 3,824 patients (Figure 1). Primary repair was most frequently performed surgical procedure in Africa reported among 1,581 (69.5%) of 2,276 patients, resection and ostomy formation in Asia reported among 563 (52.9%) of 1,065 patients, and resection and anastomosis in the Americas reported among 235 (48.7%) of 483 patients. The median year of data collection was negatively correlated with frequency of primary repair (Pearson’s rho [95% CI]: −0.39 [-0.62 - −0.11], p=<0.01). No correlation was observed between median year of data collection and frequency of resection and anastomosis, or resection and ostomy creation (Supplementary Materials Figure 4).

**Figure 4.**
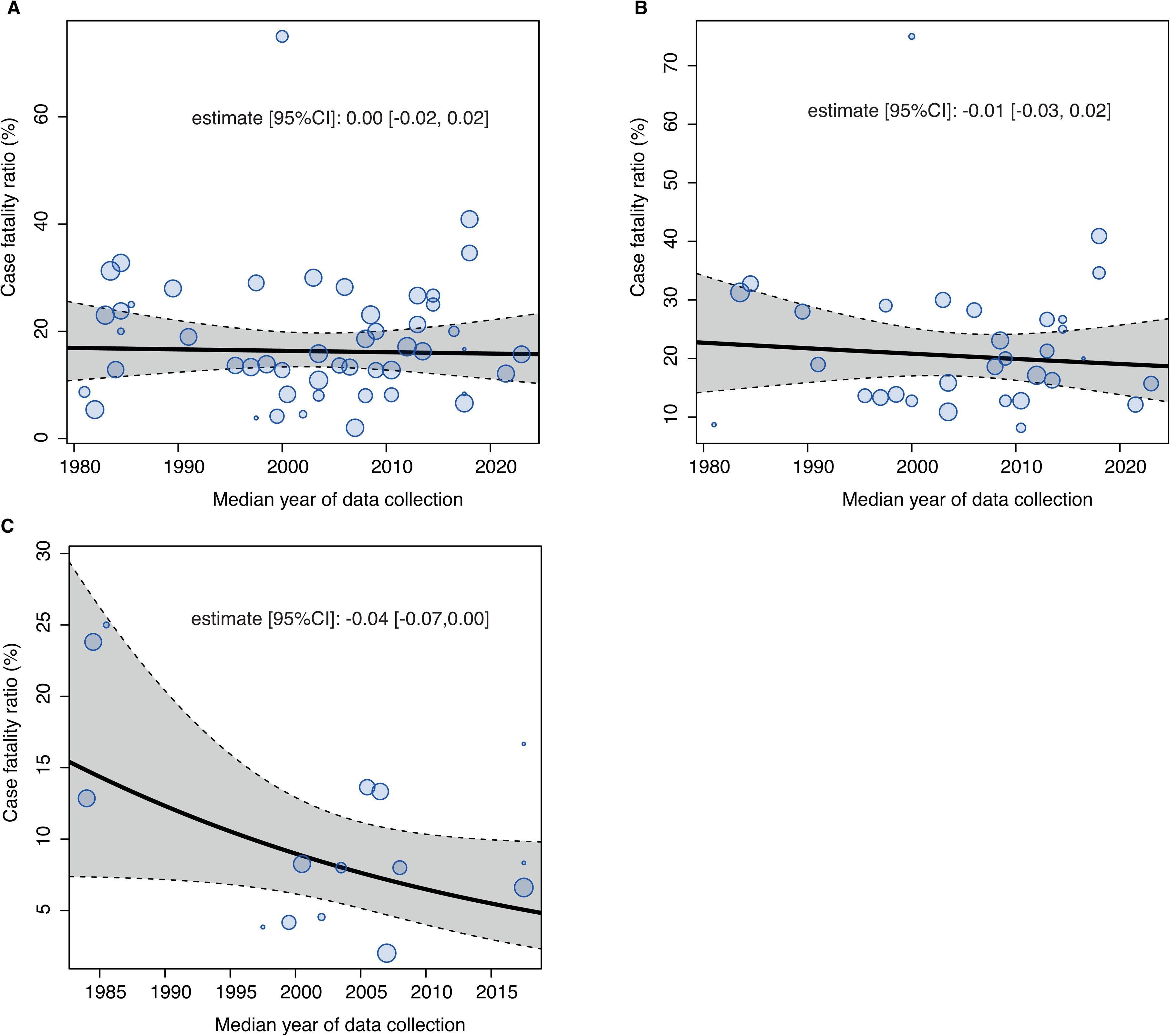
Results of meta-regression of median year of data collection with case fatality ratio of typhoid intestinal perforation, global systematic review on complications and mortality of typhoid intestinal perforation, published 1966-2023 Legend: The line shows the correlation between median year of data collection and case-fatality ratio (A, overall; B, UN region Africa; C, UN region Asia). Bubble size is proportional to study weight. Grey line represents the regression line. Shaded area bordered by dashed lines represents the 95% confidence limits.

Among 35 articles that reported any post-operative complication, the median (IQR) number of post-operative complications reported on was 7 (5-8). Of the pre-defined post-operative complications, wound or surgical site infection was the most prevalent complication occurring in 1,553 (50.1%) of 3,100 patients (Table 1). Pre-defined post-operative complications that were reported in ≥100 patients in the three UN regions were: enterocutaneous fistula, intestinal obstruction, intra-abdominal abscess, and wound or surgical site infection (Figure 2). Intestinal obstruction was reported in 15 (11.9%) of 126 patients in the Americas compared with 44 (5.6%) of 779 patients in Asia (p=0.04), and 46 (5.0%) of 911 patients in Africa (p=0.01). Wound or surgical site infections were reported in 702 (60.4%) of 1,163 patients in Asia compared with 783 (44.2%) of 1,811 patients in Africa (p<0.01). No regional differences were observed for the other comparisons, including for enterocutaneous fistula and intra-abdominal abscess. Of 27 other complications reported, complications reported in ≥five articles included sepsis in 126 (22.2%) of 567, septic shock in 47 (14.1%) of 334, and persistent peritonitis in 118 (16.7%) of 706 patients (Table 1).

**Table 1.** Post-operative complications in patients with typhoid intestinal perforations identified in the global systematic review on complications and mortality of patients with typhoid intestinal perforations, 1966-2023.

| Post-operative complication | Number of articles that reported the complication | n / N | (%) |
| --- | --- | --- | --- |
| **Pre-defined complications** |  |  |  |
| Wound or surgical site infection | 32 | 1553/3100 | (50.1) |
| Enterocutaneous fistula | 26 | 163/2531 | (6.4) |
| Intra-abdominal abscess | 24 | 238/2572 | (9.3) |
| Wound dehiscence | 17 | 308/1909 | (16.1) |
| Burst abdomen | 15 | 115/1434 | (8) |
| Intestinal obstruction | 12 | 105/1816 | (5.8) |
| Incisional hernia | 11 | 143/1522 | (9.4) |
| Respiratory infection | 11 | 136/872 | (15.6) |
| Reperforation | 10 | 115/1433 | (8) |
| Anastomotic leak | 7 | 116/1275 | (9.1) |
| **Other:** |  |  |  |
| **Cardio-respiratory** |  |  |  |
| Cardio-pulmonary complication | 3 | 14/204 | (6.9) |
| Respiratory failure | 1 | 7/12 | (58.3) |
| Septic shock | 6 | 31/235 | (13.2) |
| **Dermatologic/stoma-related** |  |  |  |
| Hypertrophic scar | 2 | 15/159 | (9.4) |
| Skin excoriation | 2 | 12/33 | (36.4) |
| Stomal prolapse | 2 | 4/33 | (12.1) |
| Stomal retraction | 3 | 6/83 | (7.2) |
| **Gastro-intestinal** |  |  |  |
| Abdominal wall fasciitis | 2 | 3/35 | (8.6) |
| Abscess buttock | 1 | 1/23 | (4.3) |
| Acute hepatic failure | 1 | 2/86 | (2.3) |
| Adhesions | 1 | 3/46 | (6.5) |
| Gallbladder disease | 1 | 5/101 | (5.0) |
| Intestinal hemorrhage | 1 | 1/5 | (20.0) |
| Persistent peritonitis | 5 | 118/706 | (16.7) |
| Prolonged ileus | 4 | 128/885 | (14.5) |
| **Hematologic** |  |  |  |
| Anemia | 2 | 14/143 | (9.8) |
| **Metabolic** |  |  |  |
| Severe electrolyte imbalance | 2 | 80/658 | (12.2) |
| **Neuropsychiatric** |  |  |  |
| Delirium | 1 | 2/25 | (8.0) |
| Psychosis | 4 | 10/254 | (3.9) |
| **Urinary** |  |  |  |
| Renal failure | 2 | 7/200 | (3.5) |
| Urinary infection | 1 | 2/45 | (4.4) |
| **Other** |  |  |  |
| Fever | 1 | 16/46 | (34.8) |
| Malnutrition | 2 | 5/24 | (20.8) |
| Multiorgan failure | 3 | 6/118 | (5.1) |
| Relapse | 1 | 1/23 | (4.3) |
| Sepsis | 9 | 126/567 | (22.2) |
| Septic arthritis | 1 | 1/50 | (2.0) |

**Table 2.**
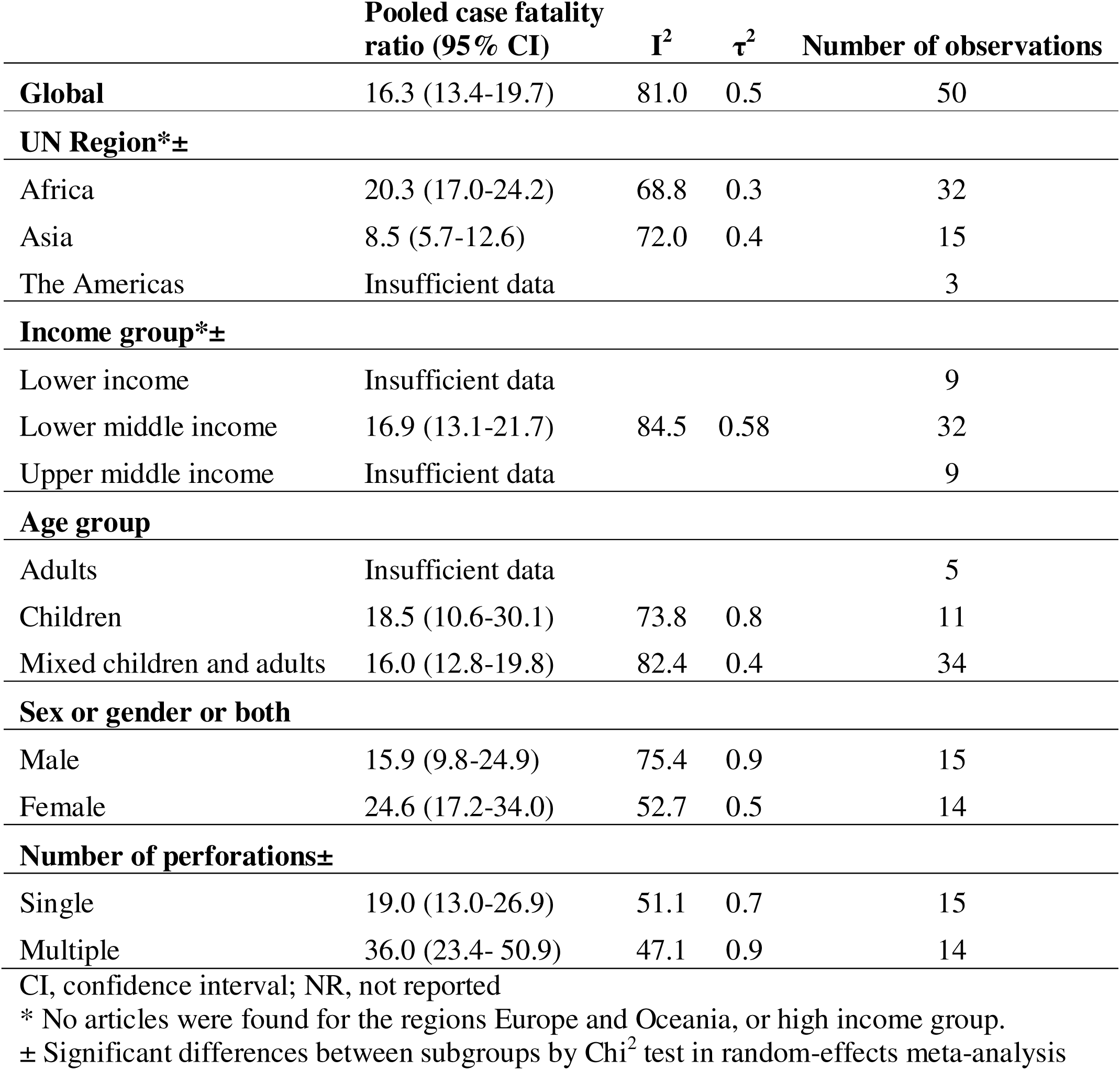
Pooled case-fatality ratio (CFR) in patients with typhoid intestinal perforations overall, per UN region, per income group, age group, sex, and number of perforations: global systematic review, 1966-2023.

The median (IQR) duration of the mean symptom onset to presentation was 11 (9-13) days in nine observations (Supplementary Materials Figure 5). The median (IQR) duration of mean symptom onset to perforation was 7 (6-12) days from five observations (Supplementary Materials Figure 6). The median (IQR) of mean length of hospital stay was 17 (12-18) days from 20 observations (Supplementary Materials Figure 7).

Among 50 observations from 47 articles reporting 583 deaths in 4,260 TIP patients, the median (IQR) CFR was 15.8% (9.2-24.7%) per estimate and the pooled CFR (95%CI) was 16.3% (13.4-19.7%). The pooled CFR in Africa was 20.3% (17.0-24.2%) and in Asia 8.5% (5.7-12.6%) (p<0.0001) (Figure 3). The pooled CFR in LMICs was 16.9% (13.1-21.7%). The pooled CFR among children was 18.5% (10.6-30.1%), and in mixed children and adults was 16.0% (12.8-19.8%). The pooled CFR among males was 15.9% (9.8-24.9%) and among females was 24.6% (17.2-34.0%) (*X*^2^, p=0.16). Insufficient estimates precluded subgroup analyses for the UN regions of the Americas and Oceania, for income groups of LIC and UIC, and for the adult age group. Pooled CFR of sex and age group by UN region are presented in Supplementary Materials 7.

Among 15 articles that reported CFR for the number of perforations, the pooled CFR (95%CI) in single perforation patients was 19.0% (13.0-26.9%), and in multiple perforation patients was 36.0% (23.4-50.9%) (*X*^2^, p<0.01). In Africa, the pooled CFR in single perforation patients was 22.0% (16.2-29.2%) and in multiple perforation patients was 42.0% (27.4-58.3%) (*X*^2^, p<0.01). Insufficient estimates precluded subgroup analyses for the UN regions of the Americas and Asia.

From meta-regression analysis among 50 observations, the median year of data collection was not correlated with the CFR (estimate [95%CI]: 0.00 [-0.02, 0.02]) (Figure 4). No correlation was observed between year of data collection and CFR in Africa (estimate [95%CI]: −0.01 [-0.03, 0.02]), and in Asia (estimate [95%CI]: −0.04 [-0.07,0.00]). Insufficient data precluded a correlation analysis for the Americas. The analyses for risk factors and moderators for CFR are presented in Supplementary Materials 8 and 9, respectively.

## Discussion

In this contemporary systematic review and meta-analysis, we estimated a TIP CFR of 16%, with higher CFR in Africa (20%) compared to Asia (9%). Over time, TIP CFR remained unchanged in Africa and Asia. Approximately 56% of patients suffered from one or more post-operative complication, of which wound or surgical site infection was the most prevalent. Higher mortality was observed in patients with multiple perforations compared to those with single perforations. Although there was a male preponderance among TIP patients, mortality between males and females was similar. For surgical procedures, regional variation was observed with primary repair most frequently performed in Africa, resection and ostomy formation most frequently reported in Asia, and resection and anastomosis in the Americas.

Consistent with the systematic review of Mogasale *et al.*,[11] our findings highlight a higher TIP CFR in Africa than in Asia and other regions. Notably, we detected an unchanged TIP CFR over time among African and Asian countries. Our findings contrast with a previous review of studies from 1990-2011 that observed a declining TIP CFR in African countries and not significant declining trend in Asian countries.[11] Besides the inclusion of more recent articles, these conflicting results could reflect the use of more robust methodology in our review including the absence of restrictions in language or country. In addition, we required that articles report data relevant to TIP case definitions to enhance attribution of perforations to *Salmonella* Typhi, and in analysis we applied the median year of data collection as a proxy for time instead of publication year.

Studies in Bangladesh,[32] India,[33] Mexico,[34, 35] Nepal,[32] Taiwan,[36] and Turkey [10, 37–39] have shown that a CFR of less than 5% is attainable for patients with TIP. The higher TIP mortality in African countries compared to Asian countries could be partially explained by a higher proportion of LICs and LMICs that showed a higher CFR in our data. In addition, data from both Africa and Asia primarily originated from university hospitals, suggesting potentially greater resources and expertise than non-university hospitals.

Availability of critical care services was only reported in 31% of articles, but was frequent in African articles. Patients with more severe disease, potentially leading to higher mortality, may be preferentially referred to hospitals with more resources. While the availability of parenteral nutrition was only mentioned in three articles, none was reported to be available in African articles. Second, the duration of symptoms before presentation and the complexity of disease, measured by multiple perforations and severe peritoneal contamination, was not significantly different between African and Asian articles. Third, primary repair was the predominant procedure in 70% of African patients, whereas resection and ostomy creation was performed in 50% of Asian patients. Although some consensus exists on the optimal surgical procedure for treating typhoid intestinal perforation,[38, 40] the choice depends on factors such as disease severity, the number and size of perforations, surgeon preference, and resource availability. Surgeons in African countries might opt for primary repair and risk an anastomotic leak or reperforation potentially avoiding complications like electrolyte disturbances and nutritional issues associated with ileostomies.

Early surgery is crucial to reduce TIP mortality.[41, 42] In the few articles with available data, we observed that duration of perforation to surgery was ≥24 hours in 63% of patients. At the article level, we observed no association between CFR and the proportion of patients with duration to surgery ≥24 hours, although data were limited. Possible reasons for surgical delays may include limited healthcare access, the logistical challenges of referral from rural hospitals that lack surgical services to those with such services, or delayed diagnosis.[43–45] Enhanced reporting and improved quality of duration data would contribute to estimating the disease burden associated with TIP.

Our comprehensive systematic review and meta-analysis involved searching multiple databases including pre-print repositories. However, there were a number of limitations. First, eligible articles were identified from a limited number of countries, reducing the generalizability of our findings to other regions. Also, the predominance of university hospitals biased our results towards hospitals equipped with more extensive surgical resources compared to lower-level hospitals. Consequently, our review has few data on hospitals with limited surgical resources, potentially leading to an underestimation of the CFR for TIP. Second, although we used a pre-defined case definition to classify patients as TIPs, only 20% of patients included in our review met the confirmed case definition.

Therefore, patients classified as probable TIP could have perforation due to non-typhoid causes. Third, most of the articles were assessed as high-risk for bias due to retrospective study designs, and a lack of clarity in the methods related to complications and routine follow up duration to ascertain complications and outcomes. Third, we reviewed potential risk factors of severe disease, but other unmeasured or unreported factors associated with severe disease such as poor access to care with surgical services could be of importance. Although some indicators of such as availability of critical care services or parenteral nutrition were poorly reported, lack of reporting does not necessarily mean lack of availability. Fourth, because our review focused on patients who underwent surgery, we did not include patients who died before reaching the hospital or surgical services. Fifth, our review lacked the ability to adjust for differences in disease severity. Last, we did not abstract data on misdiagnosis or inappropriate treatment before presentation, as our focus was on mortality, morbidity, and post-operative complications of TIP.

## Conclusion

Mortality following TIP was estimated to be 16%, with higher mortality in studies from Africa compared to those from Asia. Over time, mortality from TIP did not decline in Africa and Asia. Since morbidity and mortality associated with TIP remains substantial, implementation of efforts to reduce the occurrence of TIP, including typhoid prevention with vaccine and non-vaccine measures, early recognition and appropriate antimicrobial treatment of typhoid fever, and increased access to and quality of surgical services for those with TIP are warranted.

## Data availability statement

Underlying data are made available in an online repository https://doi.org/10.7910/DVN/JFKZOB. For additional data enquiries please contact the corresponding author.

## Ethical statement

This study was based on published data; therefore, review and approval by an institutional review board were not needed.

## Supplementary Materials

**Supplementary Materials 1**

- PRISMA checklist for the systematic review on complications and mortality of patients with typhoid intestinal perforations

**Supplementary Materials 2**

- Search strategy for the systematic review on complications and mortality of patients with typhoid intestinal perforations

**Supplementary Materials 3**

- Bias assessment methods for the systematic review on complications and mortality of patients with typhoid intestinal perforations

**Supplementary Materials 4**

- Details of included articles identified in the global systematic review on complications and mortality of patients with typhoid intestinal perforations, 1966-2023 (48 articles)

**Supplementary Materials 5**

- Descriptive characteristics by articles and by patients identified in the global systematic review on complications and mortality in patients with typhoid intestinal perforations (TIP), 1966-2023

**Supplementary Materials 6**

- Complication at presentation in patients with typhoid intestinal perforation in the global systematic review on complications and mortality of typhoid intestinal perforation, published 1966-2023, 48 articles

**Supplementary Materials 7**

- Case-fatality ratio of typhoid intestinal perforations by UN region for age group and sex, global systematic review on complications and mortality of typhoid intestinal perforation, published 1966-2023

**Supplementary Materials 8**

- Assessment of risk factors for case fatality ratio of typhoid intestinal perforation, global - systematic review on complications and mortality of typhoid intestinal perforation, published 1966-2023

**Supplementary Materials 9**

- Results of meta-regression of moderators with case fatality ratio of typhoid intestinal perforation, global systematic review on complications and mortality of typhoid intestinal perforation, published 1966-2023

**Supplementary Materials Figure 1**

- PRISMA flowchart of study selection process for the global systematic review on complications and mortality of patients with typhoid intestinal perforations, 1966-2021

**Supplementary Materials Figure 2**

- Global distribution of number of articles per country identified in the global systematic review on complications and mortality of typhoid intestinal perforations, 1966-2023 (48 articles, 51 observations)

**Supplementary Materials Figure 3**

- Frequency of risk of bias for each question, domain subtotal, and overall assessment

Legend: Details of each question are provided in Supplementary Materials 3.

**Supplementary Materials Figure 4**

- Frequency of surgical procedures by year per UN region, global systematic review on complications and mortality of typhoid intestinal perforations, 1966-2023

Legend: CI, confidence interval; UN, united nations

**Supplementary Materials Figure 5**

- Duration symptom onset to presentation in days in patients with typhoid intestinal perforation per article, systematic review, 1966-2023

Legend: SD, standard deviation

**Supplementary Materials Figure 6**

- Duration symptom onset to perforation in patients with typhoid intestinal perforation, systematic review, 1966-2023

Legend: SD, standard deviation

**Supplementary Materials Figure 7**

- Duration of hospital stay of patients with typhoid intestinal perforation per article, systematic review, 1966-2023

Legend: SD, standard deviation

