## Supplementary material for "Mortality, morbidity, and post-operative complications of typhoid intestinal perforations: global systematic review and meta-analysis"

### Supplementary Materials 1

**PRISMA checklist for the systematic review on complications and mortality of patients with typhoid intestinal perforations**

| **Topic** | **No.** | **Item** | **Location where item is reported** |
| --- | --- | --- | --- |
| **TITLE** |  |  |  |
| **Title** | 1 | Identify the report as a systematic review. | Title |
| **ABSTRACT** |  |  |  |
| **Abstract** | 2 | See the PRISMA 2020 for Abstracts checklist |  |
| **INTRODUCTION** |  |  |  |
| **Rationale** | 3 | Describe the rationale for the review in the context of existing knowledge. | Introduction |
| **Objectives** | 4 | Provide an explicit statement of the objective(s) or question(s) the review addresses. | Introduction |
| **METHODS** |  |  |  |
| **Eligibility criteria** | 5 | Specify the inclusion and exclusion criteria for the review and how studies were grouped for the syntheses. | Methods, first paragraph |
| **Information sources** | 6 | Specify all databases, registers, websites, organisations, reference lists and other sources searched or consulted to identify studies. Specify the date when each source was last searched or consulted. | Methods, first paragraph; supplemental material |
| **Search strategy** | 7 | Present the full search strategies for all databases, registers and websites, including any filters and limits used. | Supplemental material |
| **Selection process** | 8 | Specify the methods used to decide whether a study met the inclusion criteria of the review, including how many reviewers screened each record and each report retrieved, whether they worked independently, and if applicable, details of automation tools used in the process. | Paragraph 'study selection' |
| **Data collection process** | 9 | Specify the methods used to collect data from reports, including how many reviewers collected data from each report, whether they worked independently, any processes for obtaining or confirming data from study investigators, and if applicable, details of automation tools used in the process. | Paragraph 'study selection' |
| **Data items** | 10a | List and define all outcomes for which data were sought. Specify whether all results that were compatible with each outcome domain in each study were sought (e.g. for all measures, time points, analyses), and if not, the methods used to decide which results to collect. | Paragraph 'Data abstraction' |
|  | 10b | List and define all other variables for which data were sought (e.g. participant and intervention characteristics, funding sources). Describe any assumptions made about any missing or unclear information. | Paragraph 'Data abstraction' |
| **Study risk of bias assessment** | 11 | Specify the methods used to assess risk of bias in the included studies, including details of the tool(s) used, how many reviewers assessed each study and whether they worked independently, and if applicable, details of automation tools used in the process. | Paragraph 'Bias asssessment' |
| **Effect measures** | 12 | Specify for each outcome the effect measure(s) (e.g. risk ratio, mean difference) used in the synthesis or presentation of results. | Paragraph 'Data-analysis' |
| **Synthesis methods** | 13a | Describe the processes used to decide which studies were eligible for each synthesis (e.g. tabulating the study intervention characteristics and comparing against the planned groups for each synthesis (item 5)). | Paragraph 'Data-analysis' |
|  | 13b | Describe any methods required to prepare the data for presentation or synthesis, such as handling of missing summary statistics, or data conversions. | Paragraph 'Data-analysis' |
|  | 13c | Describe any methods used to tabulate or visually display results of individual studies and syntheses. | Paragraph 'Data-analysis' |
|  | 13d | Describe any methods used to synthesize results and provide a rationale for the choice(s). If meta-analysis was performed, describe the model(s), method(s) to identify the presence and extent of statistical heterogeneity, and software package(s) used. | Paragraph 'Data-analysis' |
|  | 13e | Describe any methods used to explore possible causes of heterogeneity among study results (e.g. subgroup analysis, meta-regression). | Paragraph 'Data-analysis |
|  | 13f | Describe any sensitivity analyses conducted to assess robustness of the synthesized results. | Paragraph 'Data-analysis |
| **Reporting bias assessment** | 14 | Describe any methods used to assess risk of bias due to missing results in a synthesis (arising from reporting biases). | Paragraph 'Data-analysis |
| **Certainty assessment** | 15 | Describe any methods used to assess certainty (or confidence) in the body of evidence for an outcome. | Paragraph 'Data-analysis |
| **RESULTS** |  |  |  |
| **Study selection** | 16a | Describe the results of the search and selection process, from the number of records identified in the search to the number of studies included in the review, ideally using a flow diagram. | Results first paragraph; supplemental material |
|  | 16b | Cite studies that might appear to meet the inclusion criteria, but which were excluded, and explain why they were excluded. | Not done, can be requested. |
| **Study characteristics** | 17 | Cite each included study and present its characteristics. | Supplemental material |
| **Risk of bias in studies** | 18 | Present assessments of risk of bias for each included study. | Main figure, supplemental material |
| **Results of individual studies** | 19 | For all outcomes, present, for each study: (a) summary statistics for each group (where appropriate) and (b) an effect estimate and its precision (e.g. confidence/credible interval), ideally using structured tables or plots. | Main figure |
| **Results of syntheses** | 20a | For each synthesis, briefly summarise the characteristics and risk of bias among contributing studies. | Results |
|  | 20b | Present results of all statistical syntheses conducted. If meta-analysis was done, present for each the summary estimate and its precision (e.g. confidence/credible interval) and measures of statistical heterogeneity. If comparing groups, describe the direction of the effect. | Results |
|  | 20c | Present results of all investigations of possible causes of heterogeneity among study results. | Results |
|  | 20d | Present results of all sensitivity analyses conducted to assess the robustness of the synthesized results. | Results |
| **Reporting biases** | 21 | Present assessments of risk of bias due to missing results (arising from reporting biases) for each synthesis assessed. | Results |
| **Certainty of evidence** | 22 | Present assessments of certainty (or confidence) in the body of evidence for each outcome assessed. | Results |
| **DISCUSSION** |  |  |  |
| **Discussion** | 23a | Provide a general interpretation of the results in the context of other evidence. | Discussion |
|  | 23b | Discuss any limitations of the evidence included in the review. | Discussion |
|  | 23c | Discuss any limitations of the review processes used. | Discussion |
|  | 23d | Discuss implications of the results for practice, policy, and future research. | Discussion |
| **OTHER INFORMATION** |  |  |  |
| **Registration and protocol** | 24a | Provide registration information for the review, including register name and registration number, or state that the review was not registered. | Methods |
|  | 24b | Indicate where the review protocol can be accessed, or state that a protocol was not prepared. | Methods, registry |
|  | 24c | Describe and explain any amendments to information provided at registration or in the protocol. | Not applicable |
| **Support** | 25 | Describe sources of financial or non-financial support for the review, and the role of the funders or sponsors in the review. | Funding statement |
| **Competing interests** | 26 | Declare any competing interests of review authors. | None |
| **Availability of data, code and other materials** | 27 | Report which of the following are publicly available and where they can be found: template data collection forms; data extracted from included studies; data used for all analyses; analytic code; any other materials used in the review. | Data availability statement |

### Supplementary Materials 2

**Search strategy for the systematic review on complications and mortality of patients with typhoid intestinal perforations**

**1. PubMed Search**

a. ***Search update: 01 January 2024 to 11 June 2025. Date of search: 11 June 2025***

Hits: 296

(typhi OR typhoid) AND (mortality OR "morbidity"[All Fields] OR "morbidity"[MeSH Terms] OR died or fatal* or complicat* or perforat* or bleeding or hemorr* or haemorr*) AND ("2024/01/01"[PDAT] : "3000/12/31"[PDAT]) AND hasabstract[text] ("typhi"[All Fields] OR "typhi s"[All Fields] OR ("typhoid fever"[MeSH Terms] OR ("typhoid"[All Fields] AND "fever"[All Fields]) OR "typhoid fever"[All Fields] OR "typhoid"[All Fields] OR "typhoidal"[All Fields])) AND ("mortality"[MeSH Terms] OR "mortality"[All Fields] OR "mortalities"[All Fields] OR "mortality"[MeSHSubheading] OR "morbidity"[All Fields] OR "morbidity"[MeSH Terms] OR ("death"[MeSH Terms] OR "death"[All Fields] OR "died"[All Fields]) OR "fatal*"[All Fields] OR "complicat*"[All Fields] OR "perforat*"[All Fields] OR ("bleedings"[All Fields] OR "hemorrhage"[MeSH Terms] OR "hemorrhage"[All Fields] OR "bleed"[All Fields] OR "bleeding"[All Fields] OR "bleeds"[All Fields]) OR "hemorr*"[All Fields] OR "haemorr*"[All Fields]) AND 1980/01/01:3000/12/31[Date - Publication] AND "hasabstract"[Text Word]

Translations

typhi: "typhi"[All Fields] OR "typhi's"[All Fields] typhoid: "typhoid fever"[MeSH Terms] OR ("typhoid"[All Fields] AND "fever"[All Fields]) OR "typhoid fever"[All Fields] OR "typhoid"[All Fields] OR "typhoidal"[All Fields] mortality: "mortality"[MeSH Terms] OR "mortality"[All Fields] OR "mortalities"[All Fields] OR

"mortality"[Subheading] died: "death"[MeSH Terms] OR "death"[All Fields] OR "died"[All Fields]

bleeding: "bleedings"[All Fields] OR "hemorrhage"[MeSH Terms] OR "hemorrhage"[All Fields] OR "bleed"[All Fields] OR "bleeding"[All Fields] OR "bleeds"[All Fields]

b. ***Search update: 01 January 2023 to 30 January 2024. Date of search: 30 January 2024***

Hits: 177

(((typhi OR typhoid)) AND ((mortality OR "morbidity"[All Fields] OR "morbidity"[MeSH Terms] OR died or fatal* or complicat* or perforat* or bleeding or hemorr* or haemorr*))) AND (("1980/01/01"[PDAT] : "3000/12/31"[PDAT]) AND hasabstract[text])

Translation: (("typhi"[All Fields] OR "typhi s"[All Fields] OR ("typhoid fever"[MeSH Terms] OR ("typhoid"[All Fields] AND "fever"[All Fields]) OR "typhoid fever"[All Fields] OR "typhoid"[All Fields] OR "typhoidal"[All Fields])) AND ("mortality"[MeSH Terms] OR "mortality"[All Fields] OR "mortalities"[All Fields] OR "mortality"[MeSH Subheading] OR "morbidity"[All Fields] OR "morbidity"[MeSH Terms] OR ("death"[MeSH Terms] OR "death"[All Fields] OR "died"[All Fields]) OR "fatal*"[All Fields] OR "complicat*"[All Fields] OR "perforat*"[All Fields] OR ("bleedings"[All Fields] OR "hemorrhage"[MeSH Terms] OR "hemorrhage"[All Fields] OR "bleed"[All Fields] OR "bleeding"[All Fields] OR "bleeds"[All Fields]) OR "hemorr*"[All Fields] OR "haemorr*"[All Fields]) AND (1980/01/01:3000/12/31[Date - Publication] AND "hasabstract"[Text Word])) AND (2023/1/1:2024/1/30[pdat])

c. ***Search update: 27 January 2023***

Hits: 3595

(typhi OR typhoid) AND (mortality OR "morbidity"[All Fields] OR "morbidity"[MeSH Terms] OR died or fatal* or complicat* or perforat* or bleeding or hemorr* or haemorr*) AND ("1980/01/01"[PDAT] : "3000/12/31"[PDAT]) AND hasabstract[text]

("typhi"[All Fields] OR "typhi s"[All Fields] OR ("typhoid fever"[MeSH Terms] OR ("typhoid"[All Fields] AND "fever"[All Fields]) OR "typhoid fever"[All Fields] OR "typhoid"[All Fields] OR "typhoidal"[All Fields])) AND ("mortality"[MeSH Terms] OR "mortality"[All Fields] OR "mortalities"[All Fields] OR "mortality"[MeSH Subheading] OR "morbidity"[All Fields] OR "morbidity"[MeSH Terms] OR ("death"[MeSH Terms] OR "death"[All Fields] OR "died"[All Fields]) OR "fatal*"[All Fields] OR "complicat*"[All Fields] OR "perforat*"[All Fields] OR ("bleedings"[All Fields] OR "hemorrhage"[MeSH Terms] OR "hemorrhage"[All Fields] OR "bleed"[All Fields] OR "bleeding"[All Fields] OR "bleeds"[All Fields]) OR "hemorr*"[All Fields] OR "haemorr*"[All Fields]) AND 1980/01/01:3000/12/31[Date - Publication] AND "hasabstract"[Text Word]

**Translations**

typhi: "typhi"[All Fields] OR "typhi's"[All Fields]

typhoid: "typhoid fever"[MeSH Terms] OR ("typhoid"[All Fields] AND "fever"[All Fields]) OR "typhoid fever"[All Fields] OR "typhoid"[All Fields] OR "typhoidal"[All Fields]

mortality: "mortality"[MeSH Terms] OR "mortality"[All Fields] OR "mortalities"[All Fields] OR "mortality"[Subheading]

died: "death"[MeSH Terms] OR "death"[All Fields] OR "died"[All Fields]

bleeding: "bleedings"[All Fields] OR "hemorrhage"[MeSH Terms] OR "hemorrhage"[All Fields] OR "bleed"[All Fields] OR "bleeding"[All Fields] OR "bleeds"[All Fields]

***c.	First search: 29 January 2020***

(Typhi OR typhoid) AND (mortality OR "morbidity"[All Fields] OR "morbidity"[MeSH Terms] OR died or fatal* or complicat* or perforat* or bleeding or hemorr* or haemorr*) AND ("1980/01/01"[PDAT] : "3000/12/31"[PDAT]) AND hasabstract[text]

**2. Web of Science Search**

***a. Search update: 11 June 2025***

***b. Search update: 30 January 2024***

Entitlements

- WOS: 1900 to 2024

- BCI: 1926 to 2024

- CCC: 1998 to 2024

- GRANTS: 1953 to 2024

- INSPEC: 1969 to 2024

- KJD: 1980 to 2024

- MEDLINE: 1950 to 2024

- PPRN: 1991 to 2024

- PQDT: 1637 to 2024

- SCIELO: 2002 to 2024

- ZOOREC: 1864 to 2024

| # | Search Query | Database | Results |
| --- | --- | --- | --- |
| 1 | TS=(typhi) OR TS=(typhoid) and Preprint Citation Index (Exclude – Database) | All Databases | 50855 |
| 2 | (((((((((TS=(mortality)) OR TS=(morbidity)) OR TS=(died)) OR TS=(death)) OR TS=(fatal*)) OR TS=(complicat*)) OR TS=(perforat*)) OR TS=(bleeding)) OR TS=(hemorr*)) OR TS=(haemorr) and Preprint Citation Index (Exclude – Database) | All Databases | 10112342 |
| 3 | #2 AND #1 and Preprint Citation Index (Exclude – Database) | All Databases | 10624 |
| 4 | #2 AND #1 | All Databases | 10624 |
| 5 | #2 AND #1 and 2023 or 2024 (Publication Years) | All Databases | 218 |
| 6 | #2 AND #1 and Letter or Book or Meeting or News or Editorial Material or Abstract (Exclude – Document Types) | All Databases | 9194 |
| 7 | #2 AND #1 and Letter or Book or Meeting or News or Editorial Material or Abstract (Exclude – Document Types) and Engineering or Government Law or Family Studies or Marine Freshwater Biology or Sociology or Forestry or Anthropology or Meteorology Atmospheric Sciences or Psychiatry or History Philosophy Of Science or Physics (Exclude – Research Areas) | All Databases | 8489 |
| 8 | #2 AND #1 and Letter or Book or Meeting or News or Editorial Material or Abstract (Exclude – Document Types) and Engineering or Government Law or Family Studies or Marine Freshwater Biology or Sociology or Forestry or Anthropology or Meteorology Atmospheric Sciences or Psychiatry or History Philosophy Of Science or Physics (Exclude – Research Areas) and 2023 or 2024 (Publication Years) | All Databases | 181 |

***b. Search update: 27 January 2023***

Entitlements:

- WOS: 1900 to 2023

- BCI: 1926 to 2023

- CCC: 1998 to 2023

- INSPEC: 1969 to 2023

- KJD: 1980 to 2023

- MEDLINE: 1950 to 2023

- SCIELO: 2002 to 2023

- ZOOREC: 1864 to 2023

| **#** | **Search Query** | **Database** | **Results** |
| --- | --- | --- | --- |
| #1 | TS=(typhi) OR TS=(typhoid) | All Databases | 47465 |
| #2 | (((((((((TS=(mortality)) OR TS=(morbidity)) OR TS=(died)) OR TS=(death)) OR TS=(fatal*)) OR TS=(complicat*)) OR TS=(perforat*)) OR TS=(bleeding)) OR TS=(hemorr*)) OR TS=(haemorr) | All Databases | 9008554 |
| #3 | #2 AND #1 | All Databases | 9741 |
| #4 | #2 AND #1 and Letter or Book or Meeting or News or Editorial Material or Abstract (Exclude – Document Types) and Retracted Publication or Biography (Exclude – Document Types) | All Databases | 8310 |
| #5 | #2 AND #1 and Letter or Book or Meeting or News or Editorial Material or Abstract (Exclude – Document Types) and Retracted Publication or Biography (Exclude – Document Types) and Engineering or Government Law or Family Studies or Marine Freshwater Biology or Sociology or Forestry or Anthropology or Meteorology Atmospheric Sciences or Psychiatry or History Philosophy Of Science or Physics (Exclude – Research Areas) | All Databases | 7660 |

**b.	First search: 29 January 2020**

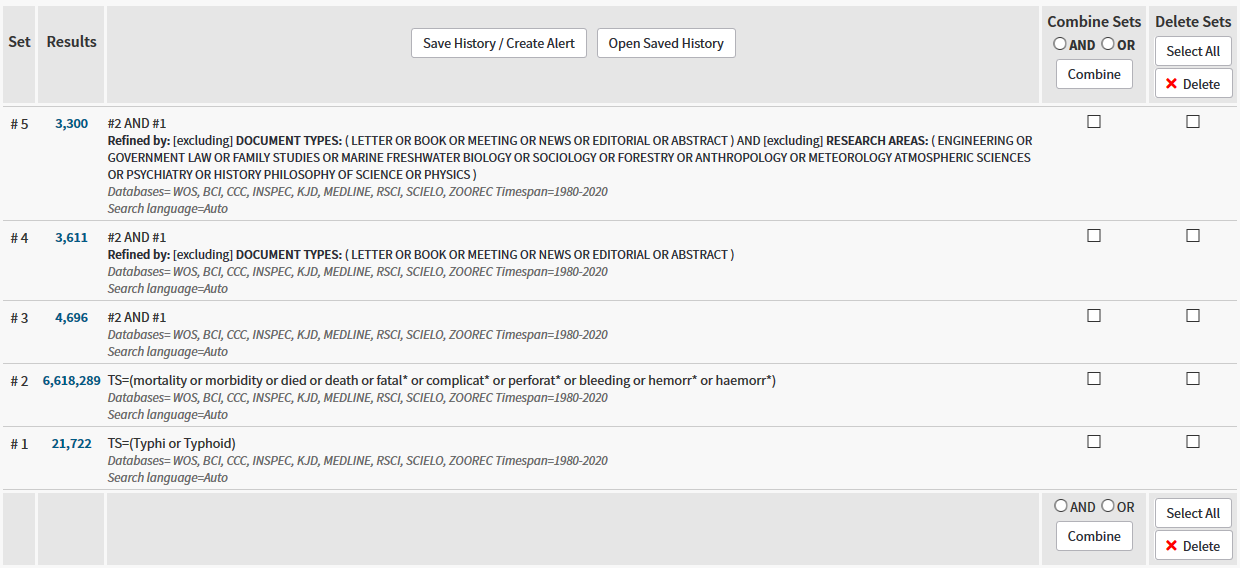

**3. Pre-print servers and pre-print repositories search: 30 January 2024, 27 January 2023, 11 June 2025**

**Database/ server**

SciLit

EuropePMC

medRxiv

NIH Preprints PMC

OSF Preprints

OpenDOAR

ROAR

JMIR Preprints

### Supplementary Materials 3

**Bias assessment methods for the systematic review on complications and mortality of patients with typhoid intestinal perforations**

| **Instrument** | **Domain [1]** | **Question** | **Levels** |
| --- | --- | --- | --- |
| JBI-prevalence | Population / setting | 1. Was the sample frame appropriate to address the target population? | yes/no/unclear/not applicable |
| JBI-prevalence | Population / setting | 2. Were study participants sampled in an appropriate way? | yes/no/unclear/not applicable |
| JBI-prevalence | Condition measurement | 6. Were valid methods used for the identification of the condition (case-definition) | yes/no/unclear/not applicable |
| JBI-prevalence | Condition measurement | 7. Was the condition measured in a standard, reliable way for all participants? | yes/no/unclear/not applicable |
| Study-specific questions | Condition measurement | Did the authors clearly define what criteria were used to clearly classify a complication? | yes/no/unclear/not applicable |
| Study-specific questions | Condition measurement | Were patients lost to follow-up and was an appropriate follow-up period defined for attributing a death to TIP? | yes/no/unclear/not applicable |
| JBI-prevalence | Statistics | 3. Was the sample size adequate? |  |
| JBI-prevalence | Statistics | 5. Was the data analysis conducted with sufficient coverage of the identified sample? | yes/no/unclear/not applicable |
| JBI-prevalence | Statistics | 8. Was there appropriate statistical analysis? | yes/no/unclear/not applicable |
| JBI-prevalence | Other | 4. Were the study subjects and the setting described in detail? | yes/no/unclear/not applicable |
| Study-specific questions | Other | What type of study design was used? | retrospective / prospective / unclear |

JBI-prevalence, Joanna Briggs institute tool for quality appraisal for prevalence studies [2]

Yes was defined as low-risk of bias; no as high-risk of bias; unclear as unclear; not applicable as not applicable

The risk of bias was summarised for each domain (population/setting; condition measurement; statistics; other) and the bias for each domain was summarised in overall risk of bias.

Calculation of domain bias:

| Domain | Domain bias level | Calculation |
| --- | --- | --- |
| Population/setting (2 items);  Statistics (3 items);  Other (2 items) | high | ≥1 items high OR 2 items unclear |
|  | intermediate | 1 item unclear |
|  | low | all others |
| Condition measurement  (4 items) | high | ≥2 items high OR ≥ items unclear |
|  | intermediate | 1 item high OR 1 item unclear |
|  | low | all others |

Calculation of overall bias:

|  | Overall bias level | Calculation |
| --- | --- | --- |
| Overall bias,  summary of four domains: | High | ≥2 domains high |
|  | Intermediate | 1 domain high |
|  | Low | all others |

### Supplementary Materials 4

**Details of included articles identified in the global systematic review on complications and mortality of patients with typhoid intestinal perforations, published 1966-2023, 48 articles**

| **First author, publication year,**  **reference** | **Country** | **Locality** | **Data collection** | **Age group** | **Quality of care** | **ICU availability** | **Overall**  **ROB** | **TIP (n)** | **Mortality TIP (n/N)** | **Male: mortality TIP (n/N)** | **Female:**  **mortality TIP (n/N)** | **Morbidity ratio (n/N)** | **Complication at presentation** | **Surgical**  **procedures** | **Post-operative complications** |
| --- | --- | --- | --- | --- | --- | --- | --- | --- | --- | --- | --- | --- | --- | --- | --- |
| Abantanga, 1997 [3] | Ghana | Kumasi | 1995-1996 | children | Antibiotics;Fluid resuscitation;Blood transfusion;Urine output monitoring;Nasogastric tube | not stated | high | 66 | 9/66 | 5/41 | 4/25 |  | *NA* | Hemicolectomy or colon resection (0/66); Other (0/66); Primary repair (66/66); Resection and anastomosis (0/66); Resection and ostomy creation (0/66); Wedge resection (0/66) | Wound or surgical site infection (32/66); Enterocutaneous fistula (1/66); Wound dehiscence (11/66); Intestinal obstruction (3/66); Burst abdomen (2/66); Incisional hernia (2/66); Chest infection (3/66) |
| Abdufatoyev, 2008 [4] | Tajikistan | Dushuanbe | 1996-2005 | children | Antibiotics;Parenteral nutrition;Fluid resuscitation;Blood transfusion;Urine output monitoring | yes | high | 109 | 9/109 |  |  | 31/109 | Gastrointestinal hemorrhage (7/109) |  | Wound or surgical site infection (11/109); Intra-abdominal abscess (5/109); Enterocutaneous fistula (2/109); Incisional hernia (15/109); Persistent peritonitis (9/109); Hypertrophic scar (3/109) |
| Adegoke, 2011 [5] | Nigeria | Ado-Ekiti (Ekiti State) | 2008-2010 | children |  | not stated | high | 47 | 6/47 |  |  |  | Anemia (8/47) |  | *NA* |
| Adesunkanmi, 1997 [6] | Nigeria | Ibadan | 1989-1990 | mixed | Antibiotics;Fluid resuscitation | not stated | high | 50 | 14/50 |  |  |  | *NA* | Hemicolectomy or colon resection (0/50); Other (0/50); Primary repair (50/50); Resection and anastomosis (0/50); Resection and ostomy creation (0/50); Wedge resection (0/50) | Wound or surgical site infection (33/50); Intra-abdominal abscess (4/50); Enterocutaneous fistula (4/50); Wound dehiscence (17/50) |
| Agu, 2014 [7] | Nigeria | Enugu | 1999-2007 | mixed | Antibiotics;Fluid resuscitation;Blood transfusion;Urine output monitoring;Nasogastric tube | not stated | high | 50 | 15/50 |  |  |  | *NA* | Hemicolectomy or colon resection (2/50); Other (0/50); Primary repair (44/50); Resection and anastomosis (4/50); Resection and ostomy creation (0/50); Wedge resection (0/50) | Wound or surgical site infection (22/50); Intra-abdominal abscess (2/50); Enterocutaneous fistula (3/50); Reperforation (3/50); Incisional hernia (8/50); Septic shock (10/50); Hypertrophic scar (12/50); Septic arthritis (1/50) |
| Ansari, 2009 [8] | Pakistan | Nawabshah | 2003-2008 | mixed | Antibiotics;Fluid resuscitation;Urine output monitoring;Nasogastric tube | not stated | high | 44 | 6/44 |  |  |  | *NA* | Hemicolectomy or colon resection (0/44); Other (0/44); Primary repair (32/44); Resection and anastomosis (4/44); Resection and ostomy creation (8/44); Wedge resection (0/44) | Wound or surgical site infection (30/44); Intra-abdominal abscess (4/44); Enterocutaneous fistula (6/44); Wound dehiscence (2/44); Incisional hernia (16/44); Stomal prolapse (3/8); Stomal retraction (2/8); Skin excoriation (8/8) |
| Atamanalp, 2007 [9] | Turkey | Erzurum | 1978-2004 | mixed | Antibiotics;Fluid resuscitation;Urine output monitoring;Nasogastric tube | not stated | high | 82 | 9/82 | 6/64 | 3/18 |  | Other shock or sepsis (27/82) | Hemicolectomy or colon resection (0/82); Other (0/82); Primary repair (32/82); Resection and anastomosis (9/82); Resection and ostomy creation (32/82); Wedge resection (9/82) | Wound or surgical site infection (45/82); Intra-abdominal abscess (6/82); Wound dehiscence (15/82); Anastomotic leak (7/82) |
| Athié, 1998 [10] | Mexico | Mexico City | 1970-1994 | mixed | Antibiotics;Parenteral nutrition | yes | high | 352 | 19/352 |  |  | 81/352 | *NA* | Hemicolectomy or colon resection (4/352); Other (0/352); Primary repair (0/352); Resection and anastomosis (226/352); Resection and ostomy creation (126/352); Wedge resection (0/352) | Enterocutaneous fistula (22/352); Anastomotic leak (51/352) |
| Birkhold, 2023 [11] | Burkina Faso; Democratic Republic Of Congo; Ethiopia; Ghana; Madagascar; Nigeria | Ouagadougo, Bale (Burkina Faso); Kisantu (DRC); Wolayita Sodo, Adama Wenji (Ethiopia); AAN & AAC, Kumasi Metropolis, Tontokrom, Keniago (Ghana); Antananarivo Renivohitra, Imerintsiatosika, Belobaka, Andina commune, Ilaka commune, Tsarasaotra commune, Antoetra commune (Madagascar); Metropolitan Ibadan, Ibarapa North (Nigeria) | 2016-2020 | mixed |  | not stated | intermediate | 225 |  |  |  |  | Gastrointestinal hemorrhage (20/225) |  | *NA* |
| Chalya, 2012 [12] | Tanzania | Mwanza | 2006-2011 | mixed | Antibiotics;Fluid resuscitation;Urine output monitoring;Nasogastric tube | yes | high | 104 | 24/104 | 17/75 | 7/29 | 41/104 | Shock (63/104) | Hemicolectomy or colon resection (8/104); Other (2/104); Primary repair (82/104); Resection and anastomosis (10/104); Resection and ostomy creation (2/104); Wedge resection (0/104) | Wound or surgical site infection (35/104); Intra-abdominal abscess (4/104); Enterocutaneous fistula (4/104); Intestinal obstruction (4/104); Burst abdomen (2/104); Incisional hernia (3/104); Chest infection (16/104); Septic shock (5/104); Prolonged ileus (2/104); Renal failure (1/104) |
| Chaudhary, 2015 [13] | India | New Delhi | 2001-2013 | mixed | Antibiotics;Fluid resuscitation | yes | high | 646 | 13/646 |  |  |  | Shock (62/646) | Hemicolectomy or colon resection (0/640); Other (0/640); Primary repair (212/640); Resection and anastomosis (24/640); Resection and ostomy creation (410/640); Wedge resection (0/640) | Wound or surgical site infection (512/633); Intra-abdominal abscess (65/633); Wound dehiscence (84/633); Intestinal obstruction (32/633); Reperforation (43/646); Incisional hernia (31/633); Anastomotic leak (48/633); Severe electrolyte imbalance (77/633); Prolonged ileus (92/633) |
| Conventi, 2018 [14] | Uganda | Karamoja | 2010-2016 | mixed | Antibiotics;Fluid resuscitation;Urine output monitoring;Nasogastric tube | not stated | high | 47 | 10/47 | 6/29 | 4/18 | 34/47 | Anemia (15/35) | Hemicolectomy or colon resection (1/45); Other (0/45); Primary repair (40/45); Resection and anastomosis (4/45); Resection and ostomy creation (0/45); Wedge resection (0/45) | Wound or surgical site infection (19/47); Enterocutaneous fistula (6/47); Wound dehiscence (13/47); Reperforation (8/47); Anastomotic leak (2/47); Septic shock (10/47); Persistent peritonitis (5/47) |
| Edino, 2004 [15] | Nigeria | Kano | 1997-2003 | mixed | Antibiotics;Fluid resuscitation;Blood transfusion | not stated | high | 47 | 6/47 |  |  |  | Gastrointestinal hemorrhage (11/47); Shock (9/47); Anemia (8/47) | Hemicolectomy or colon resection (1/47); Other (0/47); Primary repair (34/47); Resection and anastomosis (8/47); Resection and ostomy creation (0/47); Wedge resection (4/47) | Wound or surgical site infection (21/47); Intra-abdominal abscess (4/47); Enterocutaneous fistula (3/47); Wound dehiscence (15/47); Intestinal obstruction (1/47); Reperforation (0/47); Prolonged ileus (5/47) |
| Farooq, 2012 [16] | Pakistan | Lahore | 2007-2009 | mixed | Antibiotics;Fluid resuscitation;Blood transfusion;Nasogastric tube | not stated | high | 50 | 4/50 |  |  |  | Anemia (39/50) | Hemicolectomy or colon resection (0/50); Other (0/50); Primary repair (18/50); Resection and anastomosis (4/50); Resection and ostomy creation (28/50); Wedge resection (0/50) | Wound or surgical site infection (33/50); Intra-abdominal abscess (5/50); Wound dehiscence (17/50); Intestinal obstruction (3/50); Incisional hernia (5/50); Sepsis (4/50); Stomal retraction (3/50) |
| Gedik, 2008 [17] | Turkey | Diyarbakir | 1994-2005 | adults | Antibiotics;Parenteral nutrition;Fluid resuscitation;Blood transfusion | not stated | high | 96 | 4/96 | 4/83 | 0/13 | 26/96 | Psychotic states (1/96); Anemia (30/96) | Hemicolectomy or colon resection (0/96); Other (0/96); Primary repair (41/96); Resection and anastomosis (18/96); Resection and ostomy creation (37/96); Wedge resection (0/96) | Wound or surgical site infection (20/96); Intra-abdominal abscess (7/96); Intestinal obstruction (9/96); Burst abdomen (9/96); Anastomotic leak (3/96); Chest infection (9/96); Sepsis (14/96); Psychosis (1/96); Renal failure (6/96) |
| Gonzalez Ojeda, 1991 [18] | Mexico | México city | 1983-1986 | adults | Antibiotics | not stated | high | 5 | 1/5 |  |  |  | *NA* | Hemicolectomy or colon resection (0/5); Other (0/5); Primary repair (4/5); Resection and anastomosis (1/5); Resection and ostomy creation (0/5); Wedge resection (0/5) | Anastomotic leak (1/5); Septic shock (1/5); Multi organ failure (3/5); Instestinal haemorrhage (1/5) |
| Honorio-Horna, 2006 [19] | Peru | Trujillo | 1966-2000 | mixed |  | not stated | high | 126 | 29/126 | 19/97 | 10/29 | 101/126 | Gastrointestinal hemorrhage (21/126); Anemia (14/126) | Hemicolectomy or colon resection (0/126); Other (4/126); Primary repair (78/126); Resection and anastomosis (8/126); Resection and ostomy creation (36/126); Wedge resection (0/126) | Wound or surgical site infection (68/126); Intra-abdominal abscess (14/126); Enterocutaneous fistula (2/126); Intestinal obstruction (15/126); Sepsis (28/126) |
| Jeon, 2021 [20] | Madagascar | Imerintsiatosika; Antananarivo | 2016-2020 | mixed |  | not stated | high | 26 | 9/26 | 5/18 | 4/8 |  | *NA* |  | *NA* |
| Keenan, 1984 [21] | South Africa | Durban | 1979-1983 | children | Antibiotics;Fluid resuscitation | not stated | high | 23 | 2/23 |  |  | 13/23 | Anemia (12/23) | Hemicolectomy or colon resection (2/23); Other (0/23); Primary repair (19/23); Resection and anastomosis (1/23); Resection and ostomy creation (1/23); Wedge resection (0/23) | Wound or surgical site infection (7/23); Intra-abdominal abscess (1/23); Enterocutaneous fistula (2/23); Wound dehiscence (1/23); Reperforation (2/23); Chest infection (12/23); Anaemia (10/23); Abdominal wall fasciitis (1/23); Relapse (1/23); Abscess buttock (1/23) |
| Kizilcan, 1993 [22] | Turkey | Ankara | 1981-1990 | mixed | Antibiotics;Fluid resuscitation | not stated | high | 4 | 1/4 | 1/4 | 0/NA |  | Gastrointestinal hemorrhage (1/4) | Hemicolectomy or colon resection (0/4); Other (0/4); Primary repair (3/4); Resection and anastomosis (0/4); Resection and ostomy creation (1/4); Wedge resection (0/4) | *NA* |
| Kurlberg, 1991 [23] | Nepal | Kathmandu | 1981-1988 | mixed | Antibiotics;Fluid resuscitation;Urine output monitoring | not stated | high | 42 | 10/42 |  |  |  | *NA* | Hemicolectomy or colon resection (0/42); Other (0/42); Primary repair (41/42); Resection and anastomosis (1/42); Resection and ostomy creation (0/42); Wedge resection (0/42) | Wound or surgical site infection (14/42); Intra-abdominal abscess (2/42); Burst abdomen (3/42); Chest infection (5/42) |
| Mabewa, 2015 [24] | Tanzania | Mwanza | 2014-2015 | mixed | Antibiotics;Fluid resuscitation;Urine output monitoring;Nasogastric tube | not stated | low | 15 | 4/15 |  |  |  | *NA* |  | *NA* |
| Mock, 1992 [25] | Ghana | Berekum | 1978-1989 | mixed | Antibiotics | no | high | 195 | 61/195 | 38/139 | 24/56 |  | Anemia (46/142) | Hemicolectomy or colon resection (0/180); Other (0/180); Primary repair (178/180); Resection and anastomosis (2/180); Resection and ostomy creation (0/180); Wedge resection (0/180) | Wound or surgical site infection (41/195); Intra-abdominal abscess (6/195); Enterocutaneous fistula (10/195); Wound dehiscence (8/195); Intestinal obstruction (2/195); Reperforation (4/195); Chest infection (6/195) |
| Mock, 1995 [26] | Ghana | Berekum | 1990-1992 | mixed | Antibiotics;Fluid resuscitation | no | high | 58 | 11/58 |  |  |  | *NA* | Hemicolectomy or colon resection (0/58); Other (0/58); Primary repair (58/58); Resection and anastomosis (0/58); Resection and ostomy creation (0/58); Wedge resection (0/58) | *NA* |
| Ndayizeye, 2016 [27] | Rwanda | Kigali | 2014-2015 | mixed |  | yes | high | 16 | 4/16 |  |  |  | *NA* |  | *NA* |
| Nilsson, 2019 [28] | Ghana | Accra | 2009-2012 | mixed |  | not stated | high | 133 | 17/133 | 7/96 | 10/37 |  | *NA* | Hemicolectomy or colon resection (0/133); Other (0/133); Primary repair (114/133); Resection and anastomosis (19/133); Resection and ostomy creation (0/133); Wedge resection (0/133) | *NA* |
| Nuhu, 2010 [29] | Nigeria | Azare | 2004-2008 | children | Antibiotics;Fluid resuscitation;Blood transfusion | not stated | high | 46 | 13/46 |  |  | 21/46 | Anemia (37/46) | Hemicolectomy or colon resection (1/46); Other (0/46); Primary repair (38/46); Resection and anastomosis (6/46); Resection and ostomy creation (0/46); Wedge resection (1/46) | Wound or surgical site infection (21/46); Intra-abdominal abscess (1/46); Enterocutaneous fistula (4/46); Wound dehiscence (6/46); Reperforation (9/46); Chest infection (4/46); Fever (16/46); Adhesions (3/46) |
| Ogbuanya, 2023 [30] | Nigeria | Southeast | 2015-2021 | adults | Fluid resuscitation;Urine output monitoring | yes | *NA* | 44 | 18/44 |  |  |  | *NA* |  | *NA* |
| Oheneh-Yeboah, 2007 [31] | Ghana | Kumasi | 2002-2005 | adults | Antibiotics;Fluid resuscitation;Blood transfusion | not stated | high | 248 | 27/248 |  |  |  | *NA* | Hemicolectomy or colon resection (0/248); Other (0/248); Primary repair (223/248); Resection and anastomosis (25/248); Resection and ostomy creation (0/248); Wedge resection (0/248) | Wound or surgical site infection (130/248); Intra-abdominal abscess (46/248); Enterocutaneous fistula (25/248); Wound dehiscence (67/248); Intestinal obstruction (10/248); Burst abdomen (16/248); Reperforation (40/248); Incisional hernia (46/248); Persistent peritonitis (86/248) |
| Olgemoeller, 2020 [32] | Malawi | Blantyre | 2016-2017 | mixed | Antibiotics | yes | high | 10 | 2/10 | 2/NA | 0/NA |  | *NA* | Hemicolectomy or colon resection (0/10); Other (0/10); Primary repair (6/10); Resection and anastomosis (2/10); Resection and ostomy creation (2/10); Wedge resection (0/10) | *NA* |
| Osifo, 2010 [33] | Nigeria | Benin City | 1993-2007 | children | Antibiotics;Fluid resuscitation;Urine output monitoring | not stated | high | 12 | 9/12 | 6/7 | 3/5 | 12/12 | *NA* | Hemicolectomy or colon resection (0/12); Other (0/12); Primary repair (2/12); Resection and anastomosis (8/12); Resection and ostomy creation (2/12); Wedge resection (0/12) | Wound or surgical site infection (3/12); Enterocutaneous fistula (5/12); Burst abdomen (2/12); Incisional hernia (2/12); Sepsis (7/12); Psychosis (1/12); Septic shock (2/12); Malnutrition (4/12); Multi organ failure (2/12); Abdominal wall fasciitis (2/12) |
| Ouedraogo, 2017 [34] | Burkina Faso | Tenkodogo | 2010-2014 | mixed | Antibiotics | no | high | 216 | 37/216 |  |  | 156/216 | Anemia (135/216) | Hemicolectomy or colon resection (0/216); Other (0/216); Primary repair (86/216); Resection and anastomosis (98/216); Resection and ostomy creation (32/216); Wedge resection (0/216) | Wound or surgical site infection (140/216); Enterocutaneous fistula (5/216); Burst abdomen (41/216); Sepsis (62/216); Persistent peritonitis (12/216) |
| Qazi, 2020 [35] | Bangladesh; Nepal; Pakistan | Bangladesh (Dhaka); Nepal (Dhulikhel, Kathmandu); Pakistan (Karachi) | 2016-2019 | mixed | Antibiotics | not stated | intermediate | 249 | 16/249 | 9/186 | 7/63 |  | Gastrointestinal hemorrhage (17/146) |  | *NA* |
| Quealee, 2024 {Quealee, 2024 #681} | Uganda | Hoima | 2023-2023 | mixed | Antibiotics; Fluid resuscitation; Urine output; Nasogastric tube | yes | high | 70 | 11/70 |  |  | 31/70 | Anemia (70/70) |  | Wound or surgical site infection (16/70) |
| Sakiye, 2024 {Sakiye, 2024 #682} | Togo | Savannah region | 2021-2022 | children | Antibiotics; General surgeon; Fluid resuscitation | not stated | high | 99 | 12/99 |  |  | 54/99 | Shock (16/99); Anemia (26/99) | Hemicolectomy or colon resection (0/99); Other (0/99); Primary repair (30/99); Resection and anastomosis (55/99); Resection and ostomy creation (14/99); Wedge resection (0/99) | Intra-abdominal abscess (29/99); Enterocutaneous fistula (13/99); Burst abdomen (8/99) |
| Sanogo, 2013 [36] | Mali | Bamako | 2000-2007 | mixed | Antibiotics | yes | high | 120 | 19/120 |  |  |  | *NA* | Hemicolectomy or colon resection (0/120); Other (0/120); Primary repair (68/120); Resection and anastomosis (17/120); Resection and ostomy creation (35/120); Wedge resection (0/120) | Wound or surgical site infection (43/120); Enterocutaneous fistula (8/120); Burst abdomen (8/120); Anaemia (4/120); Septic shock (3/17); Cardio/pulmonary complication (10/17) |
| Sérengbé, 2002 [37] | Central African Republic | Bangui | 1997-1998 | children | Antibiotics | not stated | high | 31 | 9/31 |  |  | 14/31 | *NA* | Hemicolectomy or colon resection (0/31); Other (0/31); Primary repair (23/31); Resection and anastomosis (8/31); Resection and ostomy creation (0/31); Wedge resection (0/31) | *NA* |
| Shaikh, 2011 [38] | Pakistan | Larkana | 2006-2007 | mixed | Fluid resuscitation | not stated | high | 60 | 8/60 |  |  |  | *NA* | Hemicolectomy or colon resection (0/60); Other (0/60); Primary repair (28/60); Resection and anastomosis (8/60); Resection and ostomy creation (22/60); Wedge resection (2/60) | Wound or surgical site infection (30/60); Intra-abdominal abscess (2/60); Enterocutaneous fistula (4/60); Wound dehiscence (4/60); Anastomotic leak (4/60); Sepsis (8/8) |
| Solarana Ortiz, 2019 [39] | Angola | Huambo | 2013-2014 | mixed |  | not stated | high | 86 | 14/86 | 10/55 | 4/31 | 57/86 | *NA* | Hemicolectomy or colon resection (0/86); Other (0/86); Primary repair (54/86); Resection and anastomosis (11/86); Resection and ostomy creation (21/86); Wedge resection (0/86) | Wound or surgical site infection (31/86); Intra-abdominal abscess (10/86); Enterocutaneous fistula (1/86); Wound dehiscence (5/86); Burst abdomen (5/86); Reperforation (5/86) |
| Sümer, 2010 [40] | Turkey | Van | 1994-2010 | mixed | Antibiotics;Fluid resuscitation;Urine output monitoring;Nasogastric tube | yes | high | 22 | 1/22 |  |  | 5/22 | Gastrointestinal hemorrhage (0/22) | Hemicolectomy or colon resection (1/22); Other (0/22); Primary repair (5/22); Resection and anastomosis (16/22); Resection and ostomy creation (0/22); Wedge resection (0/22) | Wound or surgical site infection (4/22); Sepsis (1/22) |
| Tade, 2008 [41] | Nigeria | Sagamu | 1990-2004 | adults | Antibiotics | not stated | high | 105 | 14/105 |  |  | 55/105 | Gastrointestinal hemorrhage (21/105) | Hemicolectomy or colon resection (1/105); Other (0/105); Primary repair (101/105); Resection and anastomosis (3/105); Resection and ostomy creation (0/105); Wedge resection (0/105) | Wound or surgical site infection (38/105); Intra-abdominal abscess (2/105); Enterocutaneous fistula (9/105); Intestinal obstruction (12/105); Burst abdomen (4/105); Incisional hernia (8/105); Chest infection (31/105) |
| Talabi, 2014 [42] | Nigeria | Ile-Ife | 2005-2013 | children | Antibiotics;Fluid resuscitation;Blood transfusion;Urine output monitoring;Nasogastric tube | not stated | high | 45 | 9/45 | 4/26 | 5/19 | 31/36 | Anemia (5/45) | Hemicolectomy or colon resection (3/45); Other (0/45); Primary repair (37/45); Resection and anastomosis (3/45); Resection and ostomy creation (1/45); Wedge resection (1/45) | Wound or surgical site infection (29/45); Intra-abdominal abscess (2/45); Enterocutaneous fistula (4/45); Wound dehiscence (14/45); Intestinal obstruction (1/45); Burst abdomen (6/45); Reperforation (1/45) |
| Ugochukwu, 2013 [43] | Nigeria | Enugu | 2007-2009 | mixed | Antibiotics;Fluid resuscitation;Blood transfusion;Urine output monitoring;Nasogastric tube | not stated | high | 86 | 16/86 |  |  | 69/70 | Shock (77/86); Delirium or confusion (15/86) | Hemicolectomy or colon resection (9/86); Other (0/86); Primary repair (52/86); Resection and anastomosis (18/86); Resection and ostomy creation (7/86); Wedge resection (0/86) | Wound or surgical site infection (42/86); Intra-abdominal abscess (7/86); Enterocutaneous fistula (2/86); Wound dehiscence (10/86); Cardio/pulmonary complication (3/86); Persistent peritonitis (6/86); Acute hepatic failure (2/86) |
| Ugwu, 2005 [44] | Nigeria | Jos | 1994-2003 | mixed | Antibiotics;Fluid resuscitation | yes | high | 101 | 14/101 |  |  | 66/101 | Seizure or convulsions (6/101) | Hemicolectomy or colon resection (16/99); Other (0/99); Primary repair (52/99); Resection and anastomosis (31/99); Resection and ostomy creation (0/99); Wedge resection (0/99) | Wound or surgical site infection (31/101); Intra-abdominal abscess (8/101); Enterocutaneous fistula (5/101); Wound dehiscence (19/101); Intestinal obstruction (13/101); Burst abdomen (4/101); Incisional hernia (7/101); Chest infection (37/101); Psychosis (3/101); Cardio/pulmonary complication (1/101); Prolonged ileus (29/101); Gallbladder disease (5/101); Multi organ failure (1/101) |
| Usang, 2017 [45] | Nigeria | Calabar | 2006-2015 | children | Fluid resuscitation;Blood transfusion | not stated | high | 49 | 4/49 |  |  |  | Anemia (49/49) | Hemicolectomy or colon resection (2/49); Other (0/49); Primary repair (36/49); Resection and anastomosis (11/49); Resection and ostomy creation (0/49); Wedge resection (0/49) | Wound or surgical site infection (20/49); Enterocutaneous fistula (5/49); Burst abdomen (2/49); Chest infection (5/49) |
| van der Werf, 1990 [46] | Ghana | Asante-Akyim | 1982-1987 | mixed |  | not stated | high | 59 | 19/58 |  |  |  | *NA* | Hemicolectomy or colon resection (0/58); Other (0/58); Primary repair (57/58); Resection and anastomosis (1/58); Resection and ostomy creation (0/58); Wedge resection (0/58) | *NA* |
| Wabada, 2022 [47] | Nigeria | Northeastern Region | 2008-2018 | children | Antibiotics;Fluid resuscitation;Urine output monitoring;Nasogastric tube | no | high | 45 | 12/45 |  |  |  | Gastrointestinal hemorrhage (8/45); Seizure or convulsions (6/45); Anemia (21/45) | Hemicolectomy or colon resection (0/45); Other (1/45); Primary repair (31/45); Resection and anastomosis (11/45); Resection and ostomy creation (0/45); Wedge resection (0/45) | Wound or surgical site infection (29/45); Intra-abdominal abscess (2/45); Enterocutaneous fistula (8/45); Burst abdomen (3/45); Chest infection (8/45); Sepsis (1/12); Psychosis (5/45); Malnutrition (1/12); Urinary infection (2/45); Respiratory failure (7/12) |
| Waqar, 2006 [48] | Pakistan | Multan | 2003-2004 | mixed |  | not stated | high | 25 | 2/25 |  |  | 14/25 | *NA* | Hemicolectomy or colon resection (0/25); Other (0/25); Primary repair (0/25); Resection and anastomosis (0/25); Resection and ostomy creation (25/25); Wedge resection (0/25) | Wound or surgical site infection (3/25); Sepsis (1/25); Severe electrolyte imbalance (3/25); Stomal prolapse (1/25); Stomal retraction (1/25); Skin excoriation (4/25); Delirium (2/25) |

### Supplementary Materials 5

**Descriptive characteristics by articles and by patients identified in the global systematic review on complications and mortality in patients with typhoid intestinal perforations (TIP), published 1966-2023**

**Table 5.1.** Descriptive characteristics for articles by UN region

|  | **Number of articles, n=48** | | **UN region Africa, n=33** | | **UN region Asia, n=12** | | **UN region Americas, n=3** | |
| --- | --- | --- | --- | --- | --- | --- | --- | --- |
| **Article characteristics** | **n** | **%** | **n** | **%** | **n** | **%** | **n** | **%** |
| Median duration data collection in years (IQR) | 5 | (3-9) | 4 | (2-8) | 8 | (3-11) | 24 | (14-29) |
| Number of patients with TIP (IQR) | 50 | (29-100) | 50 | (45-99) | 44 | (17-83) | 126 | (66-239) |
| Number of reported surgical procedures (IQR) | 3 | (2-4) | 3 | (2-4) | 3 | (2-3) | 3 | (3-4) |
| Number of reported complications at presentation (IQR) | 1 | (1-1) | 1 | (1-1) | 1 | (1-1) | 2 | (2-2) |
| Number of reported post-operative complications (IQR) | 7 | (5-8) | 7 | (6-9) | 7 | (5-8) | 4 | (3-5) |
| *Age group* |  |  |  |  |  |  |  |  |
| Adults | 5 | 10.4 | 3 | 9.1 | 1 | 8.3 | 1 | 33.3 |
| Children | 11 | 22.9 | 10 | 30.3 | 1 | 8.3 | 0 | 0 |
| Mixed | 32 | 66.7 | 20 | 60.6 | 10 | 83.3 | 2 | 66.7 |
| *Availability of critical care services* |  |  |  |  |  |  |  |  |
| No | 4 | 8.3 | 4 | 12.1 | 0 | 0 | 0 | 0 |
| Yes | 11 | 22.9 | 7 | 21.2 | 3 | 25 | 1 | 33.3 |
| Not stated | 33 | 68.8 | 22 | 66.7 | 9 | 75 | 2 | 66.7 |
| *Hospital level** |  |  |  |  |  |  |  |  |
| District/rural | 1 | 2 | 1 | 3 | 0 | 0 | 0 | 0 |
| Regional hospital | 5 | 10 | 4 | 12.1 | 0 | 0 | 1 | 33.3 |
| University hospital | 24 | 48 | 18 | 54.5 | 5 | 35.7 | 1 | 33.3 |
| Mixed | 5 | 10 | 2 | 6 | 3 | 21.4 | 0 | 0 |
| Not stated | 15 | 30 | 8 | 24.2 | 6 | 42.9 | 1 | 33.3 |
| *Availability of care* |  |  |  |  |  |  |  |  |
| Antimicrobial treatment | 36 | 75 | 24 | 72.7 | 10 | 83.3 | 2 | 66.7 |
| Fluid resuscitation | 30 | 62.5 | 20 | 60.6 | 10 | 83.3 | 0 | 0 |
| Blood transfusion | 11 | 22.9 | 8 | 24.2 | 3 | 25 | 0 | 0 |
| Urine output monitoring | 16 | 33.3 | 11 | 33.3 | 5 | 41.7 | 0 | 0 |
| Nasogastric tubes | 13 | 27.1 | 9 | 27.3 | 4 | 33.3 | 0 | 0 |
| Parenteral nutrition | 3 | 6.3 | 0 | 0 | 2 | 16.7 | 1 | 33.3 |
| ASA classification reported for patients | 6 | 12.5 | 6 | 18.2 | 0 | 0 | 0 | 0 |

ASA, American society of anaesthesiologists; IQR, interquartile range; UN, united nations

*Based on 50 observations from 47 articles: Atamanalp et al. provided two observations for different timeperiods. Qazi et al. Provided three observations for three study sites

**Table 5.2. Descriptive characteristics of outcome variables for articles by UN region**

|  | **Number of articles, n=48** | | **Overall** | | **UN region Africa,**  **n=33** | | **UN region Asia,**  **n=12** | | **UN region Americas,**  **n=3** | |
| --- | --- | --- | --- | --- | --- | --- | --- | --- | --- | --- |
|  | N | % | Median | IQR | Median | IQR | Median | IQR | Median | IQR |
| Proportion of male# | 39 | 81.3 | 71.2 | 63.4-76.5 | 65.5 | 61.8-72.5 | 79.3 | 68.8-81.4 | 71.4 | 68.7-74.2 |
| Proportion of single perforations | 34 | 70.8 | 74.9 | 70.2-83.3 | 73.6 | 69.9-82.1 | 81.8 | 72.7-84.0 | 80 | 77.3-81.1 |
| Proportion of multiple perforations | 34 | 70.8 | 23.8 | 16.2-29.0 | 26.1 | 15.7-29.8 | 18.2 | 16.0-27.3 | 20 | 18.9-22.7 |
| Severe peritoneal contamination | 17 | 35.4 | 34.9 | 18.1-58.5 | 24.4 | 17.0-64.0 | 41.5 | 27.3-50.0 | 34.9^ |  |
| Case fatality ratio (CFR)* | 47 | 97.9 | 15.8 | 9.2-24.7 | 19.5 | 13.6-28.1 | 8 | 3.1-13.1 | 20 | 12.7-21.5 |
| CFR among adults | 5 | 10.4 | 13.3 | 10.9-20.0 | 13.3 | 12.1-27.1 | 4.2^ |  | 20^ |  |
| CFR among children | 11 | 22.9 | 13.6 | 10.4-27.5 | 16.8 | 12.3-27.9 | 8.3^ |  | ND |  |
| CFR among mixed adults/children | 31 | 64.4 | 16.1 | 9.2-23.6 | 20 | 16.1-27.3 | 8 | 2.0-13.3 | 14.2 | 9.8-18.6 |
| CFR male± | 14 | 29.2 | 18.2 | 9.0-23.8 | 20.7 | 15.4-27.3 | 4.8 | 4.8-10.7 | 19.6^ |  |
| CFR female± | 13 | 27.1 | 23.2 | 13.7-32.6 | 26.3 | 22.2-42.9 | 5.6 | 0-13.7 | 34.5^ |  |
| CFR single perforation | 15 | 31.3 | 20.6 | 15.9-26.6 | 22.6 | 17.8-28.0 | 8.9 | 5.3-17.0 | 18.1^ |  |
| CFR multiple perforations | 14 | 31.3 | 33.3 | 21.5-44.5 | 42.1 | 29.0-52.6 | 8.3 | 4.2-14.2 | 37.5^ |  |
| Morbidity ratio | 20 | 41.7 | 54.2 | 28.1-72.3 | 60.9 | 45.3-72.3 | 27.8 | 26.0-35.3 | 51.6 | 37.3-65.9 |

CFR, case-fatality ratio; IQR, interquartile range; ND, no data; UN, United Nations

*Analysis of CFR were based on 50 observations from 47 articles. Atamanalp provided two observations for different timeperiods [9] Qazi et al. Provided three observations for three study sites [35]

#Sex of patients were based on 40 observations. Atamanalp provided two observations for different timeperiods [9] Qazi et al. provided three observations for three study sites [35]

^One article

±CFR per sex was based on 15 observations. Atamanalp provided two observations for different timeperiods [9]

**Table 5.3.** **Descriptive characteristics of outcome variables by number of patients by UN region**

|  | **Overall,**  **N=4,309** | | **UN Region Africa**  **N=2,397** | | **UN Region Asia**  **N=1,429** | | **UN Region The Americas**  **N=483** | |
| --- | --- | --- | --- | --- | --- | --- | --- | --- |
|  | n/N | % | n/N | % | n/N | % | n/N | % |
| Number of patients with ASA classification |  |  |  |  |  |  |  |  |
| Class I and II | 429/746 | 57.5 | 429/746 | 57.5 | ND |  | ND |  |
| Class III and IV | 288/746 | 38.6 | 288/746 | 38.6 | ND |  | ND |  |
| Class V and VI | 29/746 | 3.9 | 29/746 | 3.9 | ND |  | ND |  |
| Proportion of male patients | 2752/4012 | 68.6 | 1440/2139 | 67.6 | 983/1404 | 70.0 | 329/478 | 68.8 |
| Proportion of single perforations | 1925/2620 | 73.5 | 1182/1645 | 71.9 | 356/492 | 72.4 | 387/483 | 80.1 |
| Proportion of multiple perforations | 695/2620 | 26.5 | 463/1645 | 28.1 | 136/492 | 27.6 | 96/483 | 19.9 |
| Severe peritoneal contamination | 523/1261 | 41.5 | 374/874 | 42.8 | 105/261 | 40.2 | 44/126 | 34.9 |
| Case fatality ratio (CFR) | 583/4260 | 13.7 | 451/2348 | 19.2 | 83/1429 | 5.8 | 49/483 | 10.1 |
| CFR in adults | 64/498 | 12.9 | 59/397 | 14.9 | 4/96 | 4.2 | 1/5 | 20 |
| CFR in children | 94/572 | 16.4 | 85/463 | 18.4 | 9/109 | 8.3 | ND |  |
| CFR in mixed adults/children | 425/3190 | 13.3 | 307/1488 | 20.6 | 70/1224 | 5.7 | 48/478 | 10 |
| CFR male | 139/920 | 15.1 | 100/486 | 20.6 | 20/337 | 5.9 | 19/97 | 19.6 |
| CFR female | 85/351 | 24.2 | 65/228 | 28.5 | 10/94 | 10.6 | 10/29 | 34.5 |
| CFR single perforation | 144/773 | 18.6 | 119/528 | 21.7 | 8/131 | 6.1 | 17/94 | 18.1 |
| CFR multiple perforations | 81/312 | 25.9 | 63/229 | 27.5 | 6/51 | 11.8 | 12/32 | 37.5 |
| Morbidity ratio | 912/1776 | 51.4 | 654/1046 | 62.5 | 76/252 | 30.2 | 182/478 | 38.1 |

ASA, American society of anaesthesiologists; CFR, case fatality ratio; ND, no data; UN, United Nations

### Supplementary Materials 6

**Complication at presentation in patients with typhoid intestinal perforation in the global systematic review on complications and mortality of typhoid intestinal perforation, published 1966-2023, 48 articles**

| **Complication**  **group** | **Complication at presentation** | **Number of patients with complication** | **% of patients with complication** | **Number of articles that reported the complication** |
| --- | --- | --- | --- | --- |
| Abdominal | Cholecystitis | ND |  |  |
|  | Gastrointestinal  haemorrhage | 106/652 | 16.3 | 9 |
|  | Hepatitis | ND |  |  |
| Cardiovascular | Asymptomatic  electrocardiographic  changes | ND |  |  |
|  | Myocarditis | ND |  |  |
|  | Shock | 227/982 | 23.1 | 5 |
| Hematologic | Anaemia | 515/1136 | 45.3 | 15 |
|  | Disseminated  intravascular coagulation | ND |  |  |
| Neuropsychiatric | Delirium or confusion | 15/86 | 17.4 | 1 |
|  | Encephalopathy | ND |  |  |
|  | Impairment of coordination | ND |  |  |
|  | Meningitis | ND |  |  |
|  | Psychotic states | 1/96 | 1.04 | 1 |
|  | Seizure or convulsions^ | 12/146 | 8.22 | 2 |
| Respiratory | Bronchitis | ND |  |  |
|  | Pneumonia | ND |  |  |
| Other | Focal abscess | ND |  |  |
|  | Pharyngitis | ND |  |  |
|  | Miscarriage | ND |  |  |
|  | Chronic carriage | ND |  |  |
|  | Shock or sepsis^ | 27/82 | 32.9 | 1 |

^: Not in predefined list

ND, no data

### Supplementary Materials 7

**Case-fatality ratio of typhoid intestinal perforations by UN region for age group and sex, global systematic review on complications and mortality of typhoid intestinal perforation, published 1966-2023**

In the African region, pooled CFR was 20.1% (11.3-33.2%) in 463 children (10 observations) and 20.9% (17.7-24.5%) for 1,488 patients of mixed children and adults (19 observations). In Africa, the CFR was 14.9% in 397 adults (three observations). In Asia, pooled CFR was 9.1% (5.7-14.3%) among 1,224 patients of mixed children and adults (13 observations). In Asia, the CFR was 8.3% in 109 children (one observation) and 4.2% in 96 adults (one observation). In the Americas, CFR from three observations was 10.1% in 483 TIP patients; of these observations, the TIP CFR was 10.1% in 478 patients of mixed children and adults (two observations) and 20.0% in five adults (one observation). Meta-analysis of TIP CFR was precluded for the Americas, for adult age group from Africa, for children and for adult age groups from Asia, and for all age groups from the Americas due to insufficient observations.

For the nine articles in the African region, the CFR was 22.7% among 486 males (nine observations) and was 16.3% among 228 females (nine observations). For five articles in Asia, CFR was 7.7% among 337 males (five observations) and was 10.6% among 94 females (four observations). Meta-analysis of TIP CFR was precluded for sex or gender or both for all UN regions due to insufficient observations. From Americas, there were 19 deaths among 97 males (one observation), and 10 deaths among 29 female (one observation).

### Supplementary Materials 8

**Assessment of risk factors for case fatality ratio of typhoid intestinal perforation, global systematic review on complications and mortality of typhoid intestinal perforation, published 1966-2023**

The median CFR was similar in 11 articles with availability of critical care facilities (15.7%) compared to four articles that reported not having critical care facilities (22.8%) (p=0.14). The median CFR was lower in three articles that reported available parenteral nutrition (5.4%) than in articles that did not (16.3%, p=0.04). No correlation was observed with CFR for the mean symptom duration until presentation (Pearson’s r: 0.33 [-0.38, 0.79], p=0.36); the proportion of patients with duration of perforation to surgery ≥24 hours (Pearson’s r: -0.30 [-0.89, 0.67], p=0.56); and the proportion of patients with severe peritoneal contamination (Pearson’s r: 0.09 [0.41-0.55], p=0.72). For hospital level, insufficient data were available for analysis.

### Supplementary Materials 9

**Results of meta-regression of moderators with case fatality ratio of typhoid intestinal perforation, global systematic review on complications and mortality of typhoid intestinal perforation, published 1966-2023**

Moderators included: country, age, study quality, income group, ICU and parenteral nutrition availability, and median year of data collection

Mixed-Effects Model (k = 50; tau^2^ estimator: Sidik Jonkman)

tau^2^ (estimated amount of residual heterogeneity): 0.3238 (SE = 0.1276)

I^2^ (residual heterogeneity / unaccounted variability): 69.56%

R^2^ (amount of heterogeneity accounted for): 35.76%

Test of moderators p-value = 0.0296

**estimate	p-value	95%	Sig#**

**confidence limits**

Intercept	66.5529	0.0946	-12.4439	145.5498	.

**Country of observation**

Bangladesh	0.3787	0.8216	-3.0630	3.8204

Burkina Faso	0.0103	0.9893	-1.5618	1.5824

Central African Republic	-0.0935	0.9249	-2.1269	1.9398

Ghana	-0.3727	0.6345	-1.9762	1.2307

India	-2.2401	0.0187	-4.0696	-0.4105	*

Madagascar	1.1542	0.1768	-0.5610	2.8694

Malawi	0.5813	0.5884	-1.6137	2.7763

Mali	-0.1439	0.8742	-2.0075	1.7197

Mexico	-1.0336	0.4250	-3.6709	1.6036

Nepal	-0.4937	0.5986	-2.4106	1.4232

Nigeria	0.0725	0.9254	-1.5157	1.6608

Pakistan	-0.6467	0.3508	-2.0535	0.7601

Peru	-0.6041	0.5341	-2.5875	1.3793

Rwanda	0.8012	0.4101	-1.1777	2.7801

South Africa A	-2.1106	0.1125	-4.7585	0.5372

Tajikistan	-0.3946	0.7646	-3.0944	2.3051

Tanzania	0.1631	0.8480	-1.5808	1.9070

Togo	-0.1386	0.8800	-2.0206	1.7434

Turkey	-1.0017	0.3121	-3.0095	1.0061

Uganda	0.4125	0.5641	-1.0483	1.8733

**Age group included**

Children	0.3846	0.3702	-0.4874	1.2566

Mixed	0.0899	0.8087	-0.6707	0.8504

**Quality of included reports**

Low	1.3041	0.0345	0.1040	2.5043	*

Moderate	0.0134	0.9837	-1.3257	1.3524

**ICU availability**

Available	-0.2283	0.5110	-0.9368	0.4803

**Parenteral nutrition availability**

Yes	-0.8830	0.2110	-2.3044	0.5384

**Median year of**	-0.0339	0.0863	-0.0731	0.0052	.

**data collection**

\# ‘Significance’ codes: 0 ‘***’ 0.001 ‘**’ 0.01 ‘*’ 0.05 ‘.’ 0.1 ‘ ’ 1

### Supplementary Materials Figure 1

**PRISMA flowchart of study selection process for the global systematic review on complications and mortality of patients with typhoid intestinal perforations, 1966-2023**

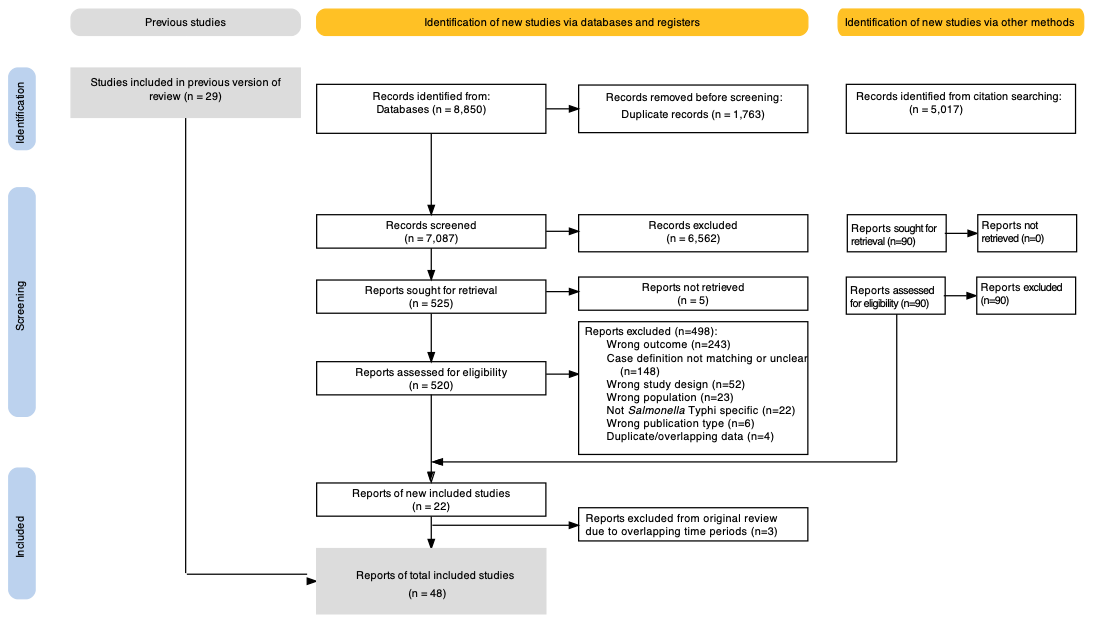

### Supplementary Materials Figure 2

**Global distribution of number of articles per country identified in the global systematic review on complications and mortality of typhoid intestinal perforations, 1966-2023 (48 articles, 51 observations)**

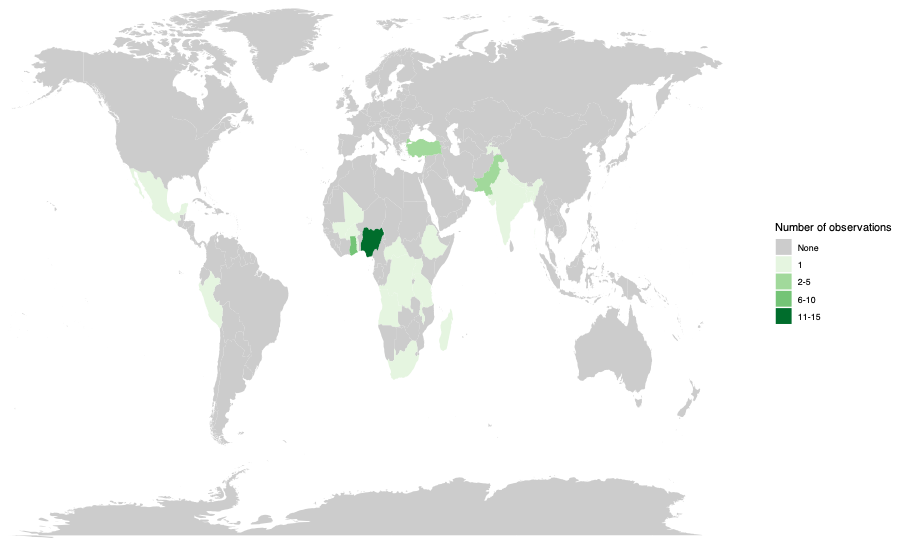

### Supplementary Materials Figure 3

**Frequency of risk of bias for each question, domain subtotal, and overall assessment**

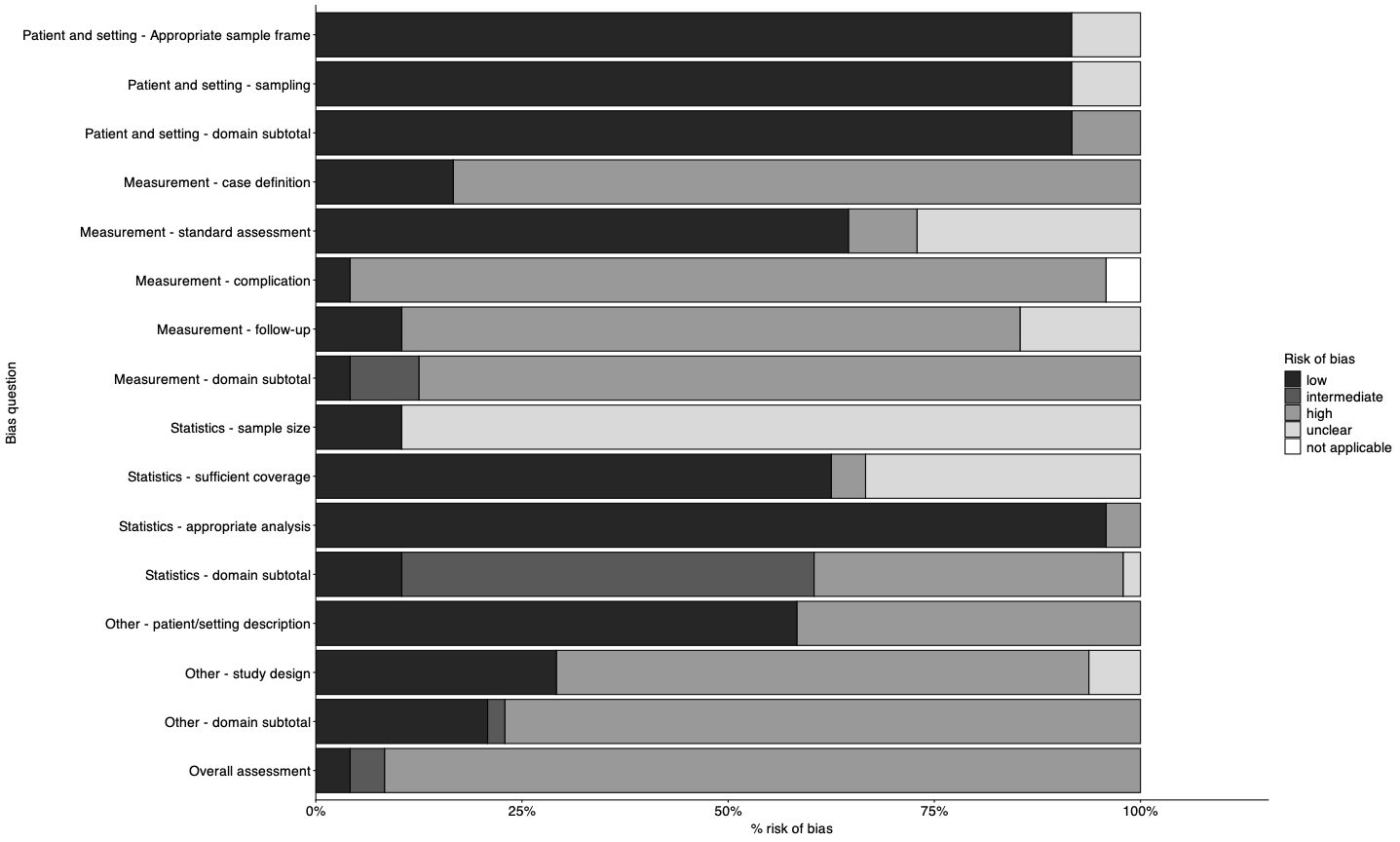

Legend: Details of each question are provided in Supplementary Materials 3.

### Supplementary Materials Figure 4

**Frequency of surgical procedures by year, global systematic review on complications and mortality of typhoid intestinal perforations, 1966-2023**

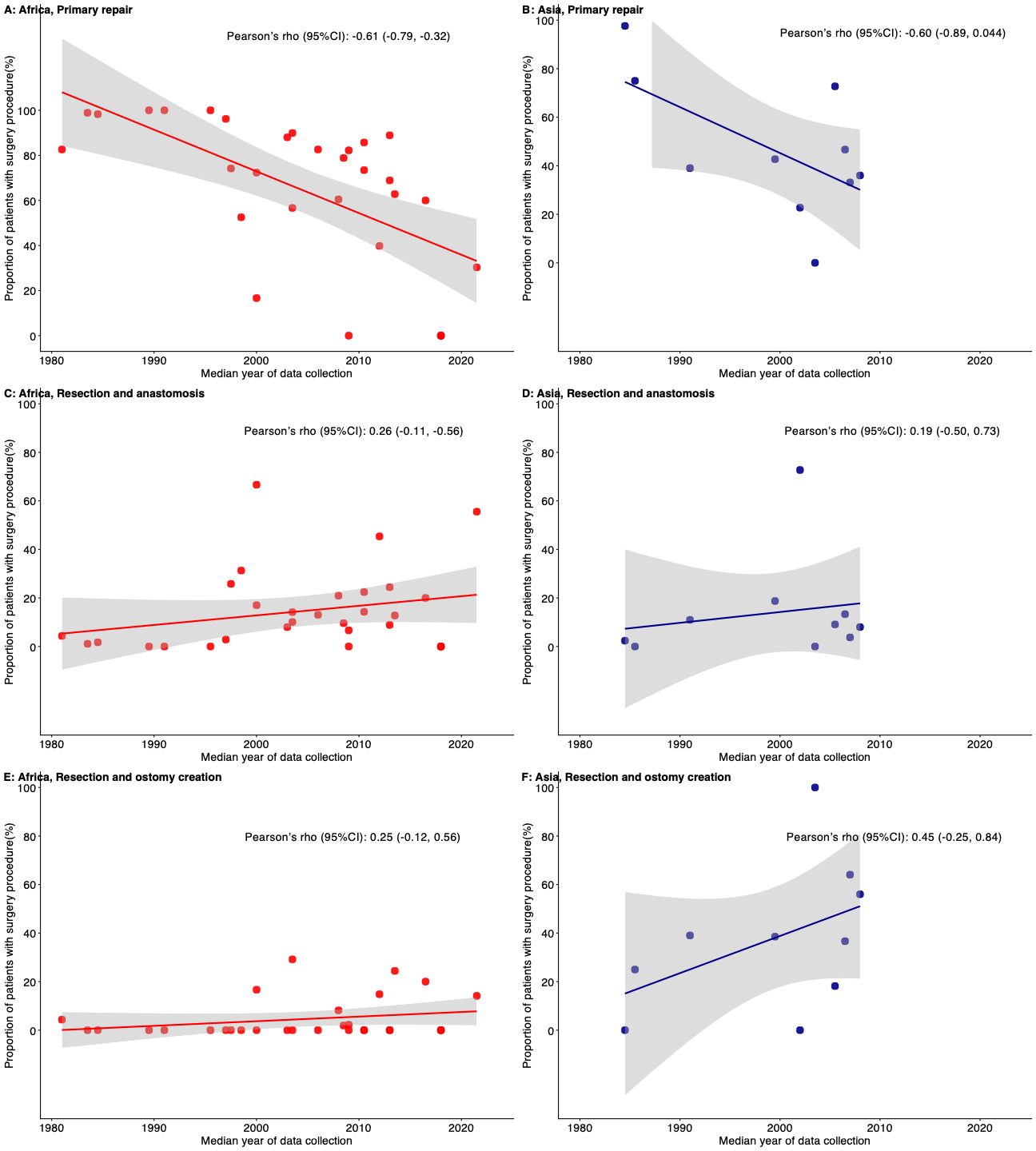

### Supplementary Materials Figure 5

**Duration symptom onset to presentation in days in patients with typhoid intestinal perforation per article, systematic review, 1966-2021**

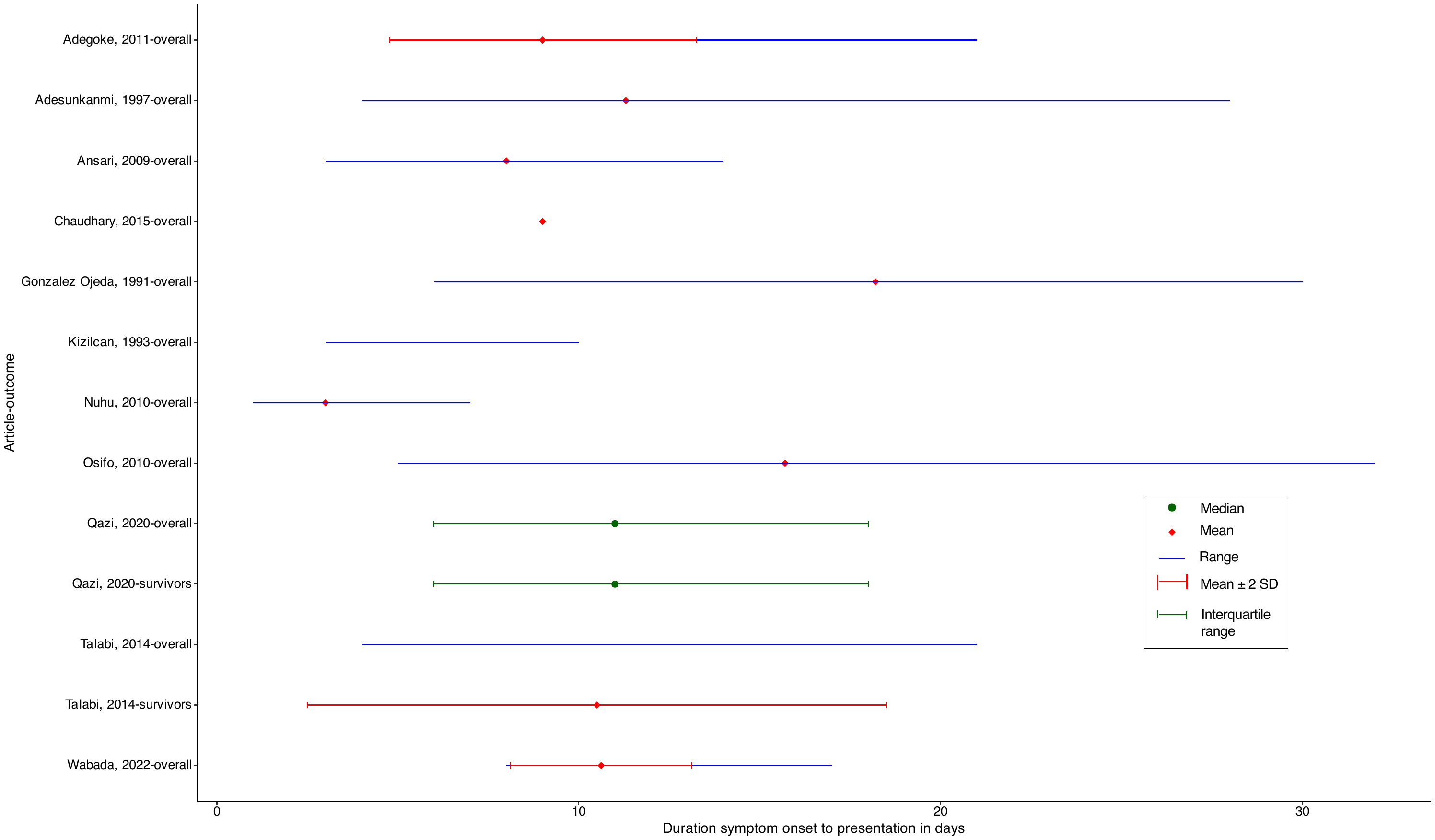

### Supplementary Materials Figure 6

**Duration symptom onset to perforation in patients with typhoid intestinal perforation, systematic review, 1966-2021**

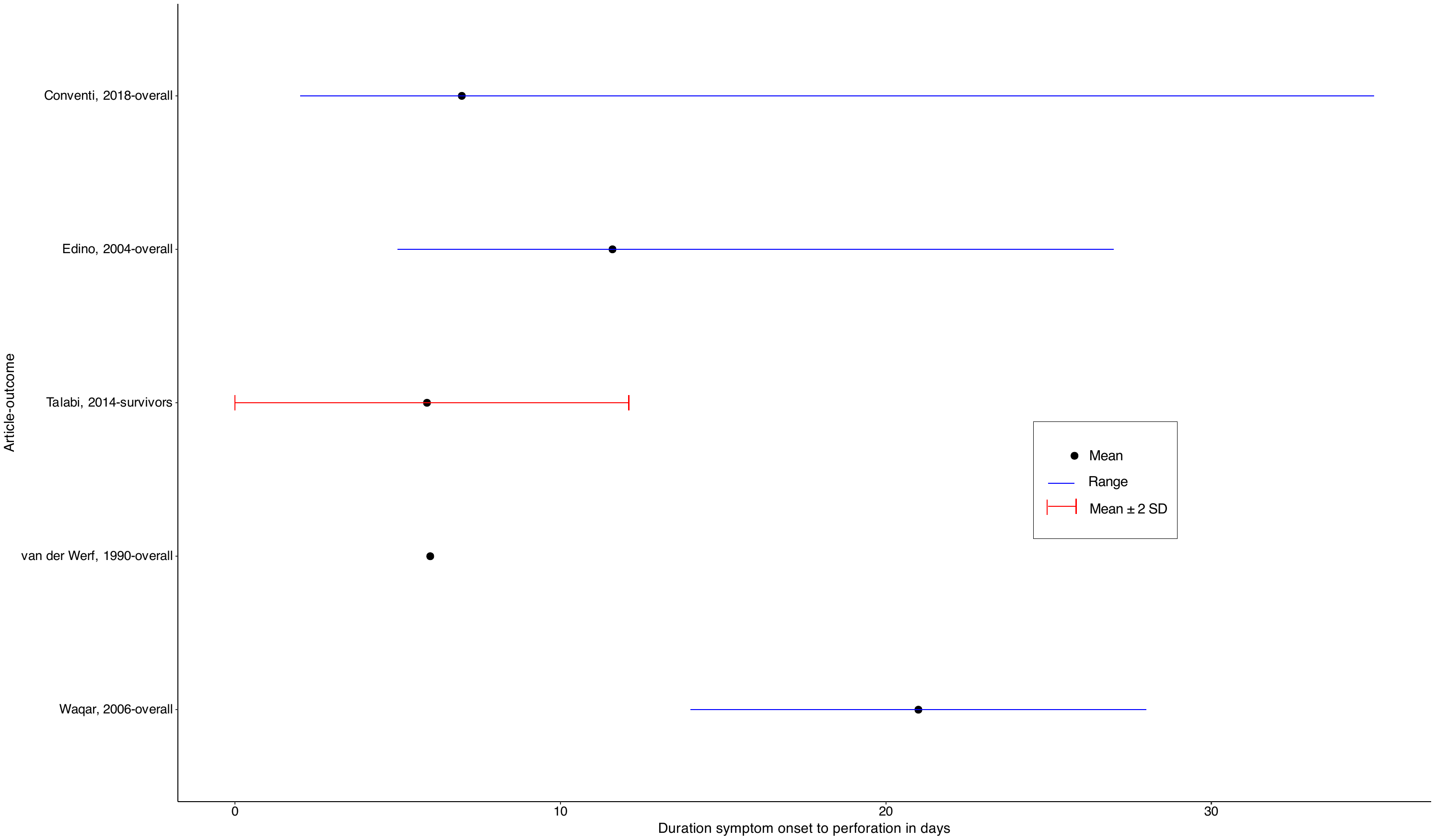

### Supplementary Materials Figure 7

**Duration of hospital stay of patients with typhoid intestinal perforation per article, systematic review, 1966-2021**

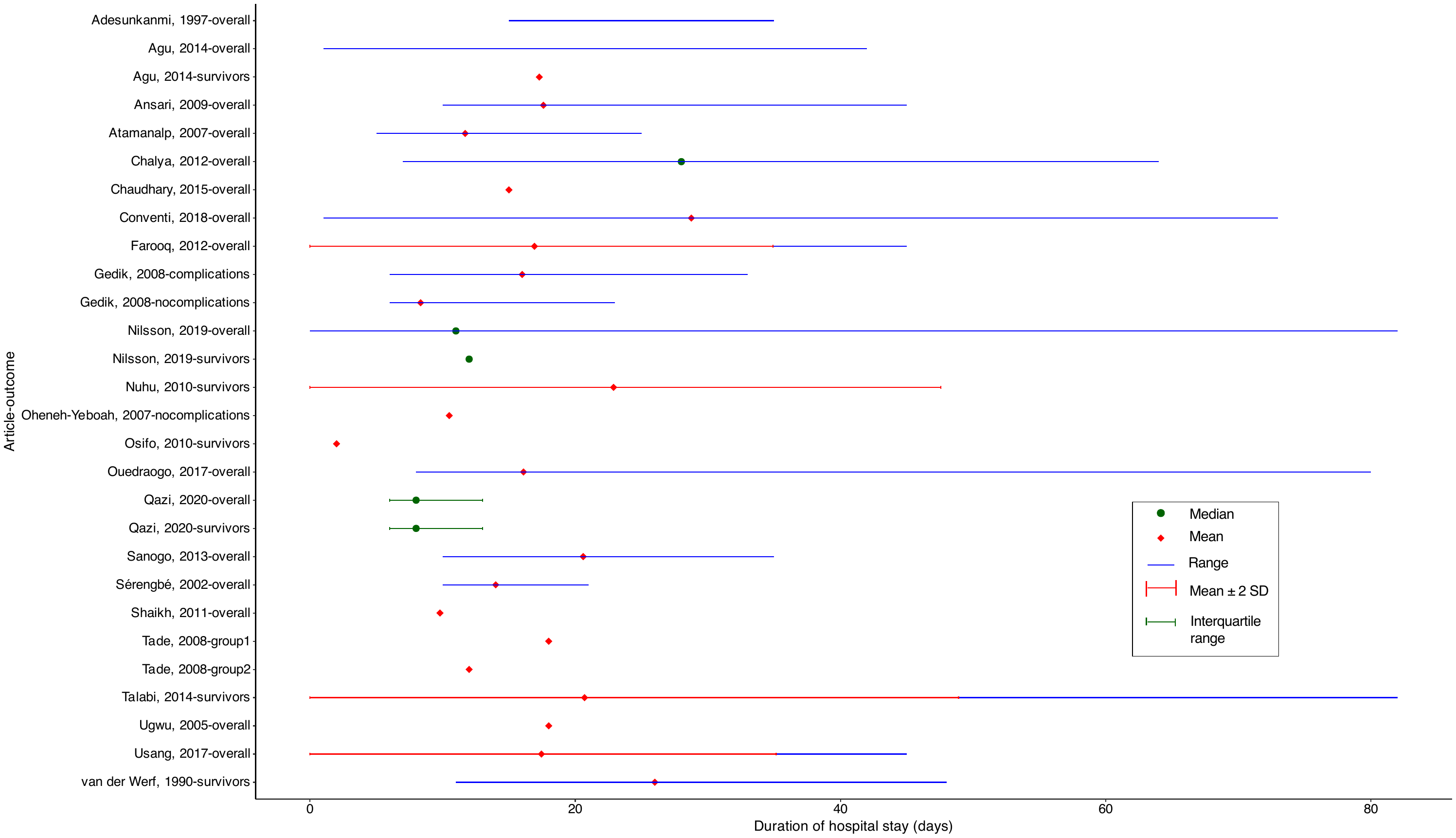
